# Polygenic prediction of treatment response for precision medicine is vulnerable to collider bias

**DOI:** 10.64898/2026.10.18.26365857

**Authors:** Matthew J Tudball, Elizabeth T Cirulli, Michael V Holmes, George Davey Smith, J Brent Richards

## Abstract

Recent reanalyses of major lipid-lowering trials have reported substantially greater treatment benefit in individuals with higher polygenic risk scores (PRSs) for coronary artery disease (CAD). We tested whether trial eligibility criteria could generate this pattern through collider bias. In simulations where the true PRS-by-treatment interaction was set to zero, we reproduced apparent PRS-dependent treatment effects similar to those observed in reanalyses of three trials. This included FOURIER-like hazard ratios for lipid-lowering treatment versus placebo of 0.77 in the high CAD PRS stratum and 0.95 in the low CAD PRS stratum. In the Helix Research Network (n = 175,106) and UK Biobank (n = 92,859) cohorts, where we did not apply the trial eligibility criteria, the CAD PRS showed little evidence of modifying the effect of statin use on acute coronary events. We then used LDL cholesterol response as a better-powered test of collider bias: we found no evidence that LDL PRS modified statin response in population cohorts, yet stratifying on baseline LDL cholesterol induced apparent PRS-dependent response matching the simulations. Importantly, treatment randomization does not protect against eligibility-induced collider bias. PRS-dependent response may therefore reflect bias from eligibility rather than biology. These results should inform the planning and execution of clinical trials for precision medicine.

**A note on this version:** Access to the UK Biobank Research Analysis Platform has been suspended since April 2026. The UK Biobank estimates reported in this article were generated before the suspension and are based on completed analyses. However, we have not been able to update Extended Data Figures 3 - 5 and Figure 8 to publication-ready versions. This limitation affects the design and presentation of the figures but does not change the underlying results. The figures will be updated once access is restored.

## Introduction

Polygenic risk scores (PRSs) are increasingly being translated into the clinic, with PRS-stratified clinical trials and patient prioritization in routine care being planned or deployed^1–3^. There are two primary reasons put forward for the use of PRS in trials. First, selecting genetically high-risk individuals for trial inclusion can increase the event rate, so-called “prognostic enrichment”, potentially shortening trial timelines and reducing the required sample size. Second, PRSs have been proposed to be able to identify individuals more likely to respond to medicines, termed “predictive enrichment”^4,5^. Here we assess whether predictive enrichment could be explained by undetected bias.

PRSs are already informing prospective trial design in anticipation that they will identify individuals more likely to benefit from therapy. The ongoing PROACT2 (NCT05850091) trial recruited individuals with high coronary artery disease (CAD) polygenic risk for randomization to rosuvastatin, colchicine, both or placebo, and the pragmatic Expression-based or Expanded PRS (EE-PRS; NCT06820086) trial randomized individuals in the highest CAD PRS stratum to rosuvastatin or usual care^2,3^. Both ongoing trials cite retrospective analyses showing greater treatment response among those with higher CAD PRS, and the EE-PRS protocol notes that lipid-lowering treatment has been shown to be “particularly effective” in people with high polygenic risk.

A major source of support for PRS-informed precision medicine comes from retrospective analyses of large lipid-lowering trials, including WOSCOPS, ODYSSEY OUTCOMES and FOURIER. These analyses report that people at higher polygenic risk of CAD receive greater benefit from pravastatin or PCSK9 inhibition than those at lower polygenic risk^6–8^. The clinically-relevant effect sizes were large. In ODYSSEY, the number needed to treat to prevent one primary endpoint was 17 in the high-PRS group compared with 67 among those at lower polygenic risk^8^. In FOURIER, the corresponding number needed to treat was 25 in the high genetic risk group compared with 143 in the low genetic risk group^6^. Because this pattern appears consistent across trials, drug classes and study populations, and because treatment was randomized, these findings have been interpreted as among the strongest evidence that PRSs can identify patients to prioritize for therapy^4,5,9^.

However, randomization only balances treatment arms among those who enter a trial; it does not make trial eligibility independent of genetic risk, which can introduce collider bias. Collider bias is a systematic distortion of relationships among variables that jointly cause selection into a population^10,11^. When genetic risk and other risk factors both affect trial eligibility, they can become spuriously associated with each other within the trial population, even if no such associations exist in the broader population. For example, FOURIER and ODYSSEY enrolled patients with established cardiovascular disease and persistently elevated LDL cholesterol despite statin therapy. A patient with a high CAD PRS may satisfy these criteria partly because of their pre-existing genetic risk, whereas a patient with a low CAD PRS may require a greater burden of other risk factors to become eligible. Conversely, a patient with a low burden of other risk factors may need elevated genetic risk to meet the eligibility criteria. Consequently, within the selected trial population, CAD PRS can become negatively correlated with other determinants of CAD risk or, in other words, lower genetic risk tends to imply greater burden of other risk factors, and vice versa.

Importantly, we identified that the trial reanalyses exhibited this negative association between CAD PRS and other risk factors (**Results**). Therefore, within the trial population, a higher CAD PRS can become a proxy for lower burden of other CAD risk factors (**Fig. 1**). Even if CAD PRS does not modify treatment response to PCSK9 inhibition, it may, through associations induced by collider bias, proxy other factors that do modify treatment response. Therefore, apparent comparatively beneficial effects of treatment in individuals at a high CAD PRS may be entirely a consequence of trial selection, rather than biologic effects of the PRS (**Fig. 2**)^10,12^. Crucially, although randomization of treatment preserves balance of these factors between treatment arms, it does not prevent this collider association from biasing the estimated PRS-by-treatment interaction.

**Figure 1:**
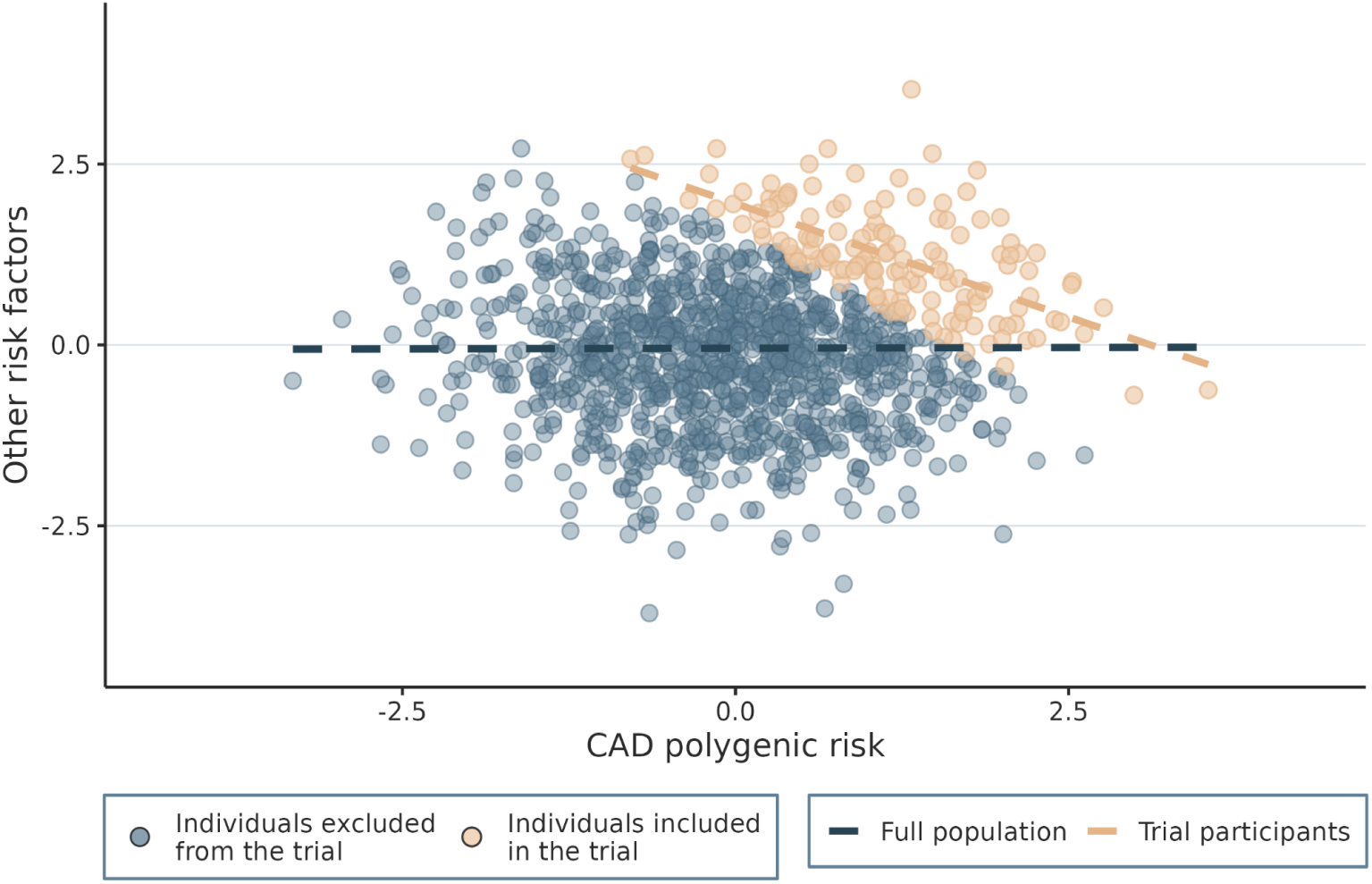
In an illustrative example depicted in Figure 1, a clinical trial recruits patients with established CAD. Both a high CAD PRS and high levels of other risk factors contribute to CAD, so trial eligibility can induce a negative association between them among trial participants, even when they are independent in the source population. The blue dashed line represents the correlation between CAD PRS and other risk factors in the general population, which is null; the red dashed line represents the same correlation among trial participants, which is strongly negative.

**Figure 2:**
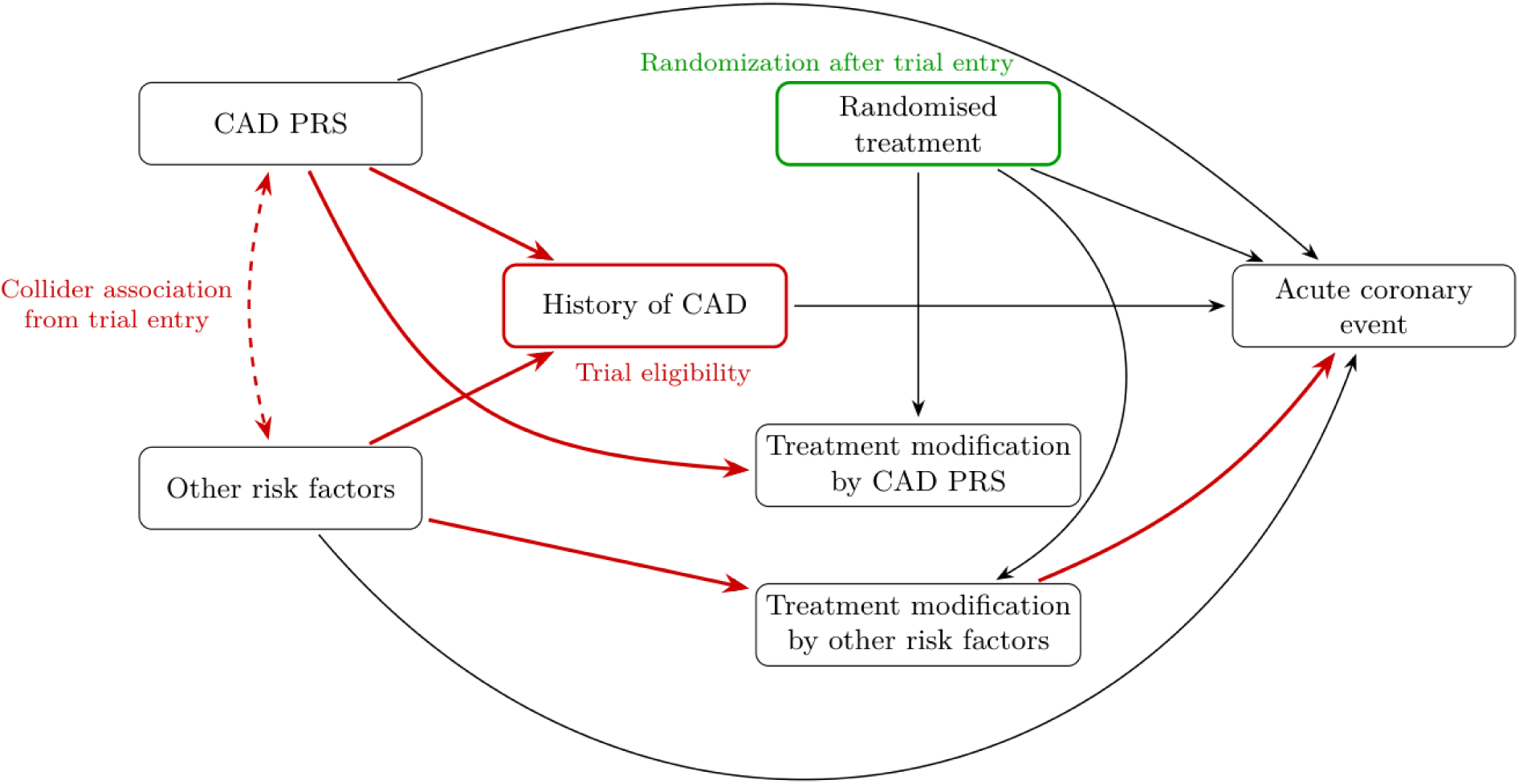
In this directed acyclic graph (DAG), entry into a clinical trial depends on both CAD PRS and other risk factors, since both influence the development of CAD. As a result, among individuals who are eligible for the trial, CAD PRS and other risk factors become correlated. Because other risk factors may modify how patients respond to lipid-lowering therapy, this induced relationship can make CAD polygenic risk appear to modify treatment effects on acute coronary events, even when no true biological interaction exists. The pathways contributing to this bias are highlighted in red. Randomization balances baseline characteristics between treatment arms, but does not remove the association between CAD PRS and other risk factors introduced by the trial eligibility criteria.

A similar concern can arise in observational data such as biobanks and electronic health records. These cohorts are usually more representative and thus less prone to eligibility-induced collider bias, but there is no treatment randomization, making them susceptible to confounding if analysis methods are not structured carefully. In observational studies, PRS-by-treatment interactions can therefore be confounded by the determinants of treatment prescribing and adherence. Analyses often address this confounding by adjusting for, stratifying on, or restricting by likely confounders, such as baseline LDL cholesterol.

However, as demonstrated previously using UK Biobank data, restricting analyses to biomarker values or disease histories can distort analyses in a similar way to trial eligibility^13^. Reflecting this tension, methodological recommendations in pharmacogenetics remain inconsistent, with some arguing against baseline adjustment and others recommending it^14,15^. Inverse probability of treatment weighting (IPTW) within a target trial emulation framework^16,11,17^ was designed to handle variables like baseline LDL cholesterol that can act as both confounders and colliders^18,19^. By appropriately reweighting individuals to balance LDL cholesterol and other baseline measurements between treatment initiators and non-initiators, IPTW aims to create quasi-randomized treatment assignment analogous to that of a clinical trial, while also retaining the representativeness of routine-care data. This balancing means that variables like pre-treatment LDL cholesterol should not confound estimates of treatment modification by PRS, and the lack of adjustment or stratification prevents the introduction of collider bias.

Here, we test whether collider bias may explain the association of a CAD PRS with outcomes in FOURIER, ODYSSEY and WOSCOPS PRS re-analyses using simulations, causal diagrams and real-world observational data analyses of statin response. In simulations with no true PRS-dependent treatment response by design, we observed the qualitative and, in many cases, quantitative findings of the three major trial reanalyses. This occurred despite complete randomization of treatment and despite the trials using different eligibility criteria, including requiring recent coronary disease in an ODYSSEY-like simulation and excluding prior myocardial infarction in a WOSCOPS-like simulation. We next tested the most direct analogue of these findings in the Helix Research Network (HRN; N = 175,106) and UK Biobank primary care records (UKB; N = 92,859), where we found little evidence that CAD PRS modified the effect of statin therapy on acute coronary events^20^. Because these analyses were limited by relatively few clinical events, we next used LDL cholesterol response as a better-powered test of the proposed collider mechanism (HRN, N = 78,145; UKB, N = 28,299). LDL PRS showed little evidence of modifying statin response in the full cohorts, whereas stratifying the same populations on baseline LDL cholesterol induced apparent PRS-dependent treatment response consistent with the simulation predictions.

Importantly, our findings do not undermine the use of PRSs for prognostic enrichment, where identifying individuals at increased disease risk can increase trial event rates and improve trial efficiency^4,5^. Rather, they provide an alternative explanation for the widely-reported effects of response to therapies stratified by polygenic risk. Such observed effects may result from inclusion criteria which induce collider bias. These findings challenge a central assumption in genomic precision medicine: observed responses to therapy, stratified by genetic risk, may not arise from biological effects, but rather from bias. In the example of a CAD PRS, apparent genetic modification of lipid-lowering therapy can arise from trial eligibility and study design choices, even when there is no effect of the PRS on treatment response. These findings are relevant not only to the design of clinical trials, but also for the translation of PRSs into clinical practice.

## Results

### Published trial populations show inverse associations between CAD PRS and other CAD risk factors

We first examined whether the published trial populations from FOURIER, ODYSSEY and WOSCOPS showed a pattern that would be expected due to eligibility-induced collider bias. In UK Biobank, CAD polygenic risk was not associated with age, smoking or diabetes in the general population, but became significantly and negatively associated with these factors among individuals with prevalent CAD^13^. This means that selecting patients with CAD, as was done in the published trials, could induce a negative correlation between CAD PRS and other CAD risk factors. We identified this same pattern in the actual published retrospective analyses of FOURIER, ODYSSEY and WOSCOPS. In FOURIER, a higher CAD PRS was associated with lower prevalence of diabetes (P = 0.007), lower prevalence of smoking (P = 0.01) and younger age (P < 0.0001)^6^. ODYSSEY showed a similar association with younger age (P < 0.0001) and WOSCOPS with lower prevalence of smoking (P = 0.006)^7,8^.

These findings are consistent with the pattern expected if trial eligibility induces an inverse relationship between genetic risk and other determinants of cardiovascular disease. They do not establish that collider bias caused the reported PRS-dependent treatment effects, but they do demonstrate that an observable feature predicted by this mechanism was present in the same trial populations in which those effects were reported. We therefore next tested whether trial eligibility capable of generating these associations could also produce apparent PRS-dependent treatment effects.

### Simulations without a PRS-by-treatment interaction recapitulate FOURIER-, ODYSSEY- and WOSCOPS-like findings

We developed a single simulation framework that could be adapted to the eligibility criteria of FOURIER, ODYSSEY and WOSCOPS (**Methods**; **Extended Data Fig. 1**). In the simulated source population, CAD polygenic risk and a single variable representing an aggregate of other CAD risk factors were chosen to be independent, but both contributed to LDL cholesterol and cardiovascular disease risk, which, in turn, determined trial eligibility. The other risk factor also modified treatment response, whereas CAD PRS did not. This means that, by design, we would not expect CAD PRS to predict treatment response in the tested population, however, if such findings were observed, they could be due to bias.

Simulation parameters were anchored to published or observed quantities wherever possible, including LDL cholesterol distributions, genetic associations, treatment effects, event rates, trial sample sizes and eligibility thresholds; for less directly estimable parameters, we specified plausible ranges and examined their influence in sensitivity analyses (**Tables 1**–**2**). We then imposed trial-specific combinations of LDL cholesterol level and cardiovascular disease history, with treatment randomized only after trial entry. Thus, a high PRS or a high burden of other risk factors could act as alternative routes into the trial, allowing eligibility to associate CAD PRS with a true treatment response modifier via collider bias. Our objective was to assess whether, in a simulated setting with a fixed ground truth of no PRS-dependent treatment response, collider bias was sufficient to reproduce the FOURIER, ODYSSEY and WOSCOPS findings.

**Table 1:**
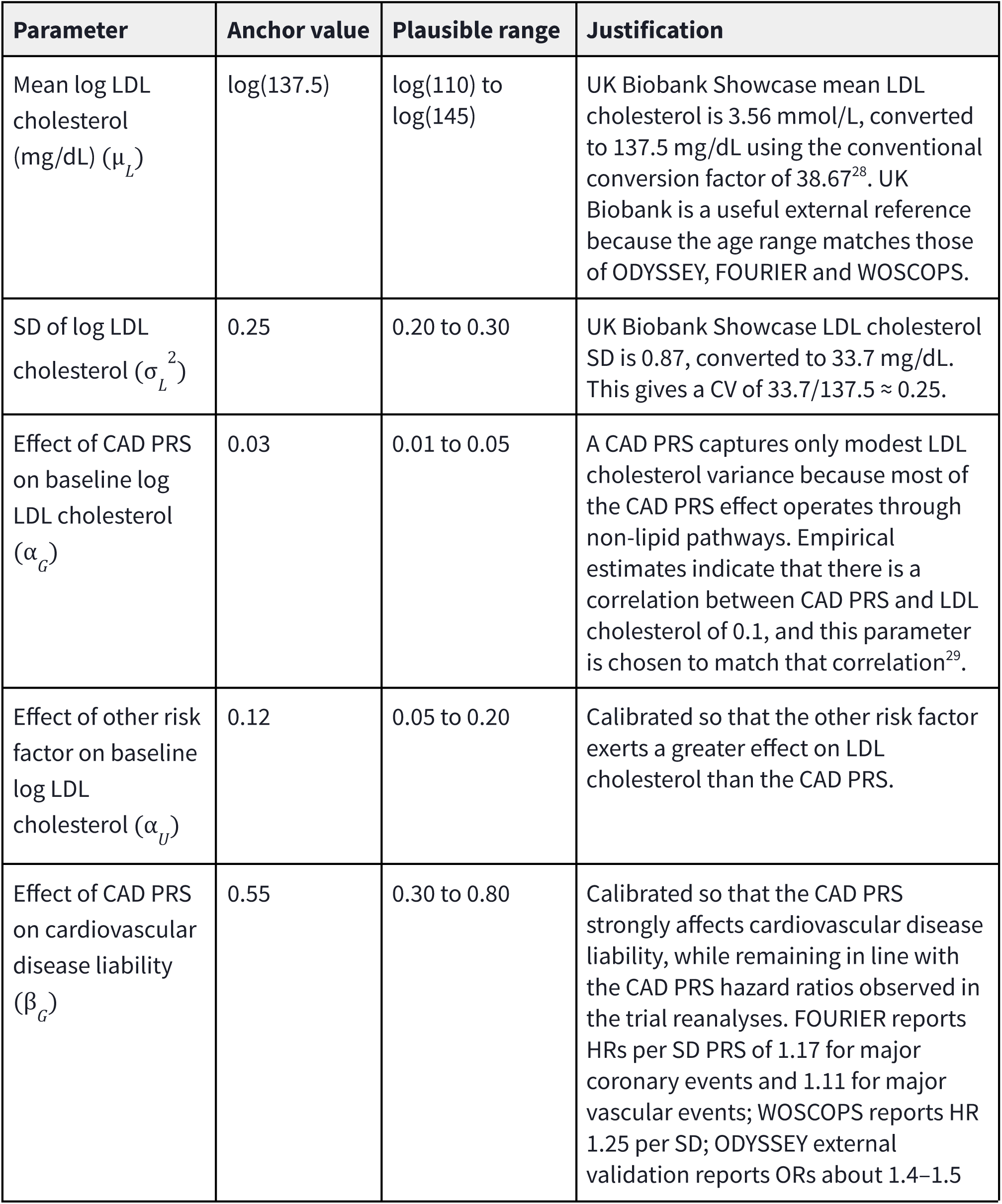
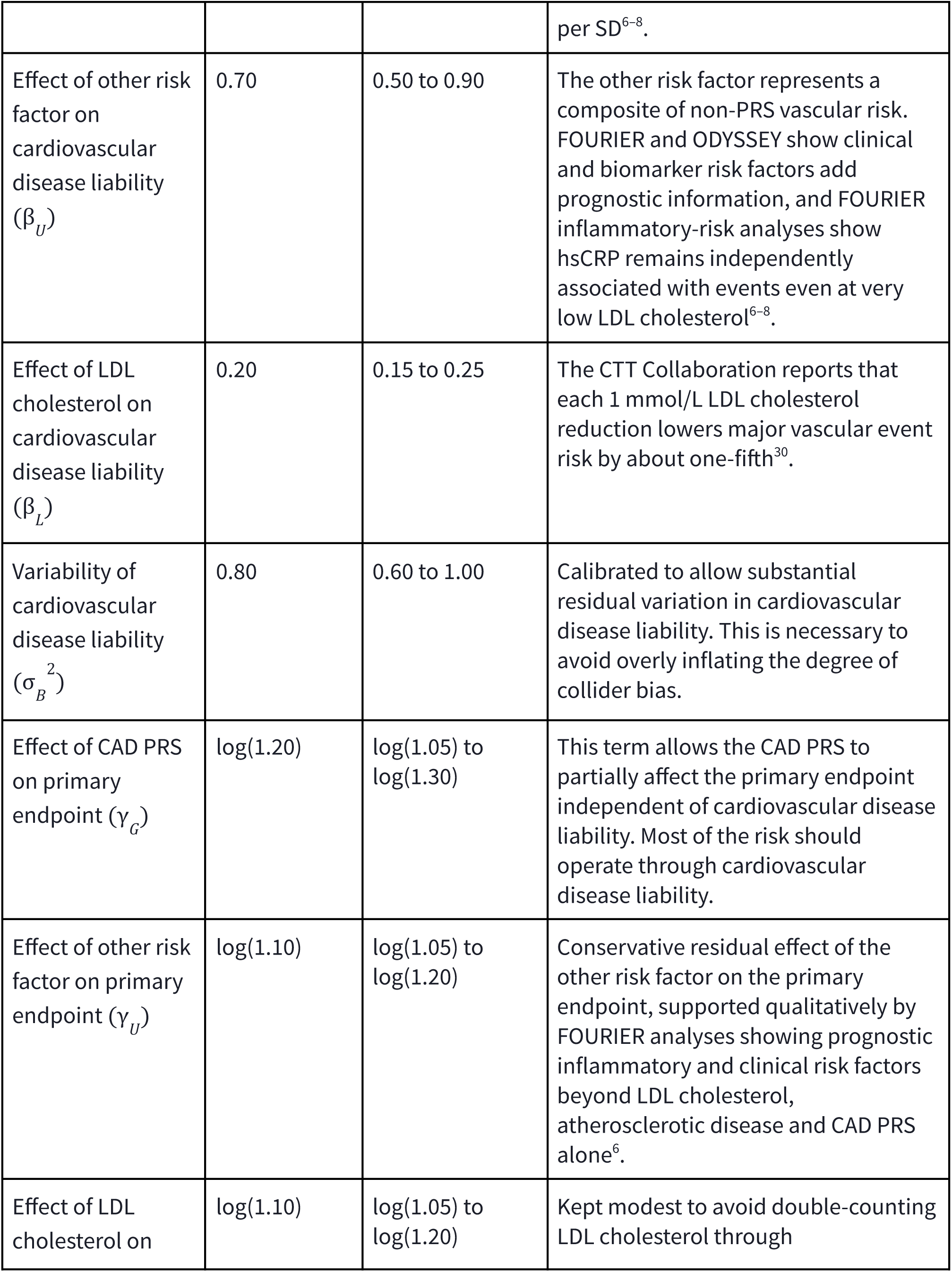
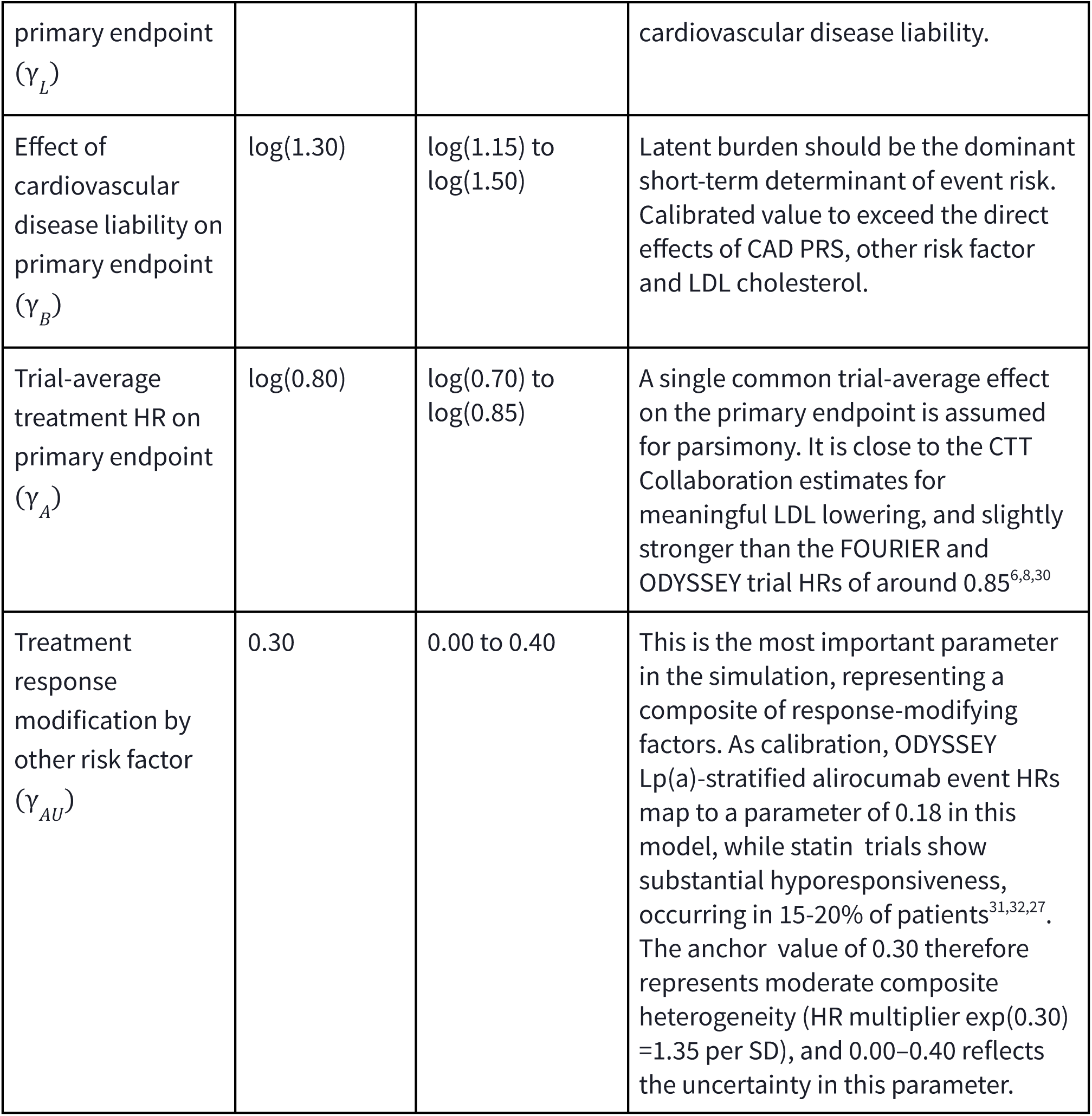
Fixed parameters common across FOURIER, ODYSSEY and WOSCOPS trial simulations.

**Table 2:** Trial-specific parameters chosen to represent the essential but simplified characteristics of FOURIER, ODYSSEY and WOSCOPS trials.

| Trial | Parameter | Anchor value | Justification |
| --- | --- | --- | --- |
| FOURIER | Trial size / treated / placebo | 14,298 / 7,163 / 7,135 | Matches the FOURIER genetic subset<sup>6</sup>. |
| FOURIER | Placebo event rate | 0.115 | FOURIER genetic reanalysis placebo major vascular event rates were 9.8%, 11.5% and 13.2% across low/intermediate/high PRS groups. The anchor value is the midpoint<sup>6</sup>. |
| FOURIER | Minimum LDL threshold ($\tau_L$) | 130 mg/dL | FOURIER required LDL cholesterol ≥ 70 mg/dL on statin therapy. With 69% high-intensity and 30% moderate-intensity statin use, the untreated equivalent cut-off is approximately 130 mg/dL<sup>6</sup>. |
| FOURIER | Percentile of cardiovascular disease liability corresponding to established ASCVD prevalence ($p$) | 0.05 | Parsimonious proxy for established ASCVD: prior MI, nonhemorrhagic stroke or symptomatic PAD. |
| ODYSSEY | Trial size / treated / placebo | 11,953 / 5,967 / 5,986 | Matches the ODYSSEY pharmacogenomic analysis cohort<sup>8</sup>. |
| ODYSSEY | Placebo event rate | 0.120 | ODYSSEY placebo MACE incidence was 17% in the top PRS decile and 11.4% in the lower 90%, with a weighted average of around 12%<sup>8</sup>. |
| ODYSSEY | Minimum LDL threshold ($\tau_L$) | 140 mg/dL | ODYSSEY required recent ACS and LDL cholesterol ≥ 70 mg/dL, non-HDL-C ≥ 100 mg/dL, or ApoB ≥ 80 mg/dL despite intensive/maximally-tolerated statin therapy. With 88.8% high-intensity statin use, the untreated-equivalent LDL cholesterol threshold is about 140 mg/dL<sup>8</sup>. |
| ODYSSEY | Percentile of cardiovascular disease liability corresponding to recent ACS ($p$) | 0.02 | Captures sharper selection for recent ACS 1-12 months before randomisation, rather than stable ASCVD in FOURIER<sup>8</sup>. |
| WOSCOP<br>S | Trial size / treated /<br>placebo | 4,892 / 2,452 /<br>2,440 | Approximate WOSCOPS genetic<br>subset<sup>7</sup>. |
| WOSCOP<br>S | Placebo event rate | 0.142 | WOSCOPS low-PRS placebo event<br>rate was 12.9% and high-PRS placebo<br>event rate was 19.6%, with a<br>weighted average of around 14.2%<sup>7</sup>. |
| WOSCOP<br>S | Minimum LDL threshold<br>($\tau_L$) | 174 mg/dL | WOSCOPS enrolled men aged 45-64<br>years with LDL cholesterol $\geq 4.5$<br>mmol/L, approximately 174 mg/dL<sup>7</sup>. |
| WOSCOP<br>S | Percentile of<br>cardiovascular disease<br>liability corresponding<br>to prior MI / clinical<br>CHD among high LDL<br>cholesterol individuals<br>($p$) | 0.05 | Parsimonious proxy for WOSCOPS<br>exclusion of prior MI/clinical CHD<br>among high LDL cholesterol<br>individuals<sup>7</sup>. |

### Bias from trial eligibility can reproduce FOURIER-, ODYSSEY- and WOSCOPS-like findings

In the simulation, we reproduced the same pattern of increased apparent treatment benefit among high-PRS individuals. This occurred despite treatment being randomly assigned within each simulated trial and despite the outcome model containing no true effect of the PRS on treatment response. PRS subgroup comparisons were structured to match the corresponding retrospective trial reanalyses. Each trial simulation was run 1,000 times at the corresponding trial sample size, and results were averaged across the simulations.

In the FOURIER-like simulation, the mean treatment hazard ratio for major vascular events was 0.95 in the low PRS group, 0.84 in the intermediate group and 0.77 in the high PRS group, closely mirroring the real trial findings (**Fig. 3**). The ODYSSEY-like simulation produced a similar pattern, with a mean hazard ratio for major adverse cardiovascular events of 0.75 in the high PRS group and 0.85 among all other participants (**Fig. 3**). The WOSCOPS-like simulation was less closely aligned with the real trial findings, but still showed greater apparent benefit in the high PRS group, with a mean hazard ratio for incident coronary heart disease of 0.81 compared with 0.88 among all other participants (**Fig. 3**).

**Figure 3:**
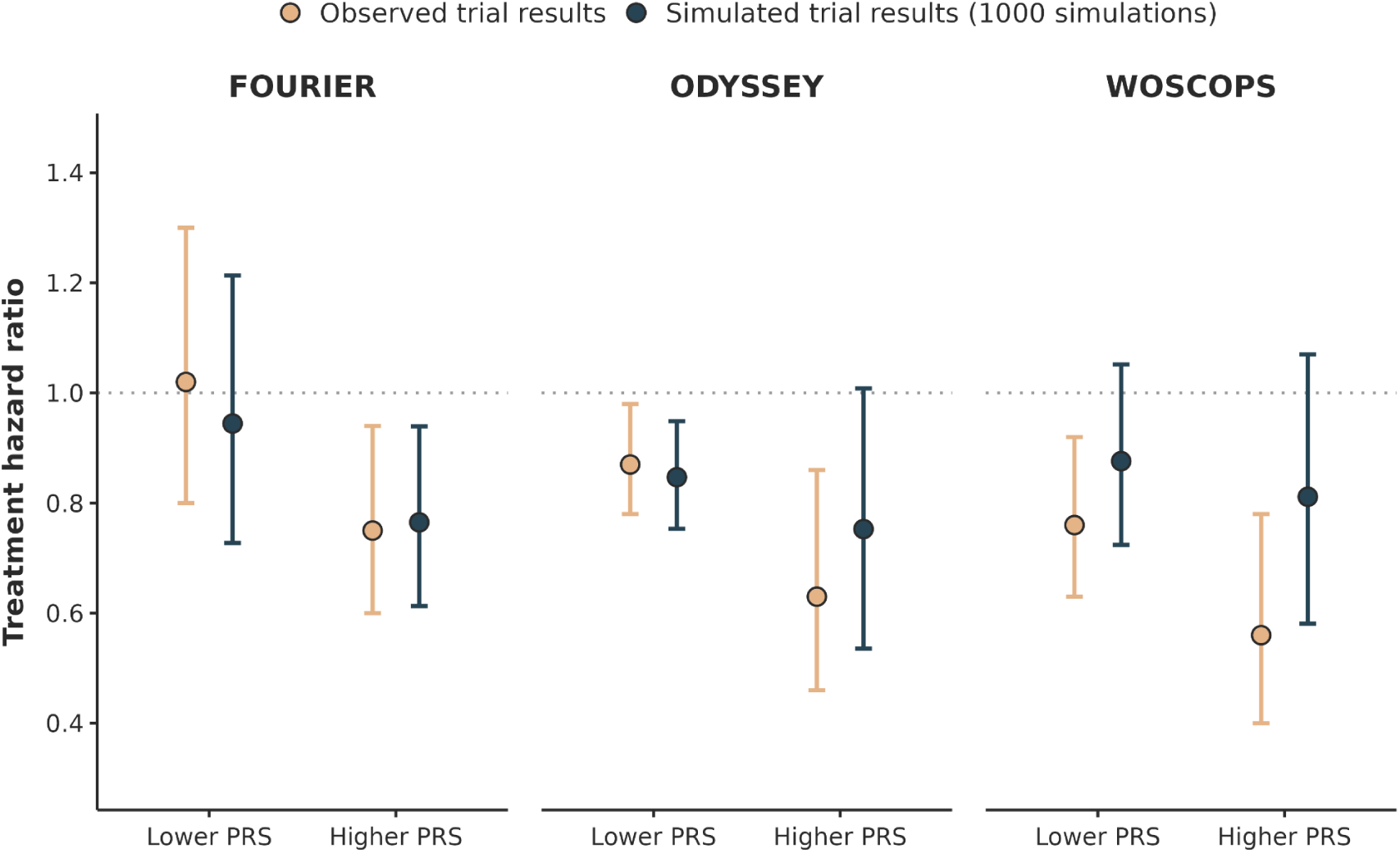
Published and simulated subgroup treatment hazard ratios across FOURIER-, ODYSSEY- and WOSCOPS-like analyses. Published subgroup hazard ratios are shown in red and simulated hazard ratios in blue. Simulated estimates are averaged across 1,000 simulated trials generated at the corresponding trial sample size; error bars show the 2.5th–97.5th percentile range across simulations. The simulation was generated from data where there was no interaction between the PRS and treatment effect. Trial-specific eligibility criteria was applied. Values further below 1 indicate greater apparent treatment benefit.

Although the simulation replicated the qualitative pattern of increased treatment response in the high-PRS group despite designing the simulation such that there were no true treatment response effects of the PRS, we also asked how often the simulation replicated the magnitude of the difference across PRS groups observed in the published trial results. To do this, we calculated the percentage of 1,000 simulations with a high-versus-lower PRS treatment HR ratio lying within the 95% confidence interval of the published HR ratio. Such HR ratio magnitudes occurred in 91% of FOURIER-like simulations, 81% of ODYSSEY-like simulations and 79% of WOSCOPS-like simulations (**Extended Data Fig. 2**). Thus, in simulations capturing the core eligibility structure of each trial, collider bias from trial eligibility was sufficient to reproduce both the pattern and magnitude of apparent PRS-related treatment benefit in the majority of simulations across all three trials. The trial reanalyses had wide confidence intervals for these HR ratios, so the high degree of compatibility reflects, in part, large statistical uncertainty as well.

The key parameter in our simulation is the extent to which the other CAD risk factor modifies response to treatment. We therefore tested how the percentage of simulations lying inside the 95% confidence intervals of the published trial results changes as this parameter is varied (**Methods**; **Fig. 4**). When this parameter is zero, there is no collider bias and the simulations fall within the published 95% confidence intervals 52-68% of the time, which is reflective of relatively high statistical uncertainty in the trial reanalyses’ estimates. As treatment modification from the other CAD risk factor increased, compatibility with the published findings also increased, reaching 95% for FOURIER, 87% for ODYSSEY and 85% for WOSCOPS at a maximum value of 0.40. These findings are consistent with the hypothesis that at least some of the observed effect of the high PRS on treatment response was due to collider bias.

**Figure 4:**
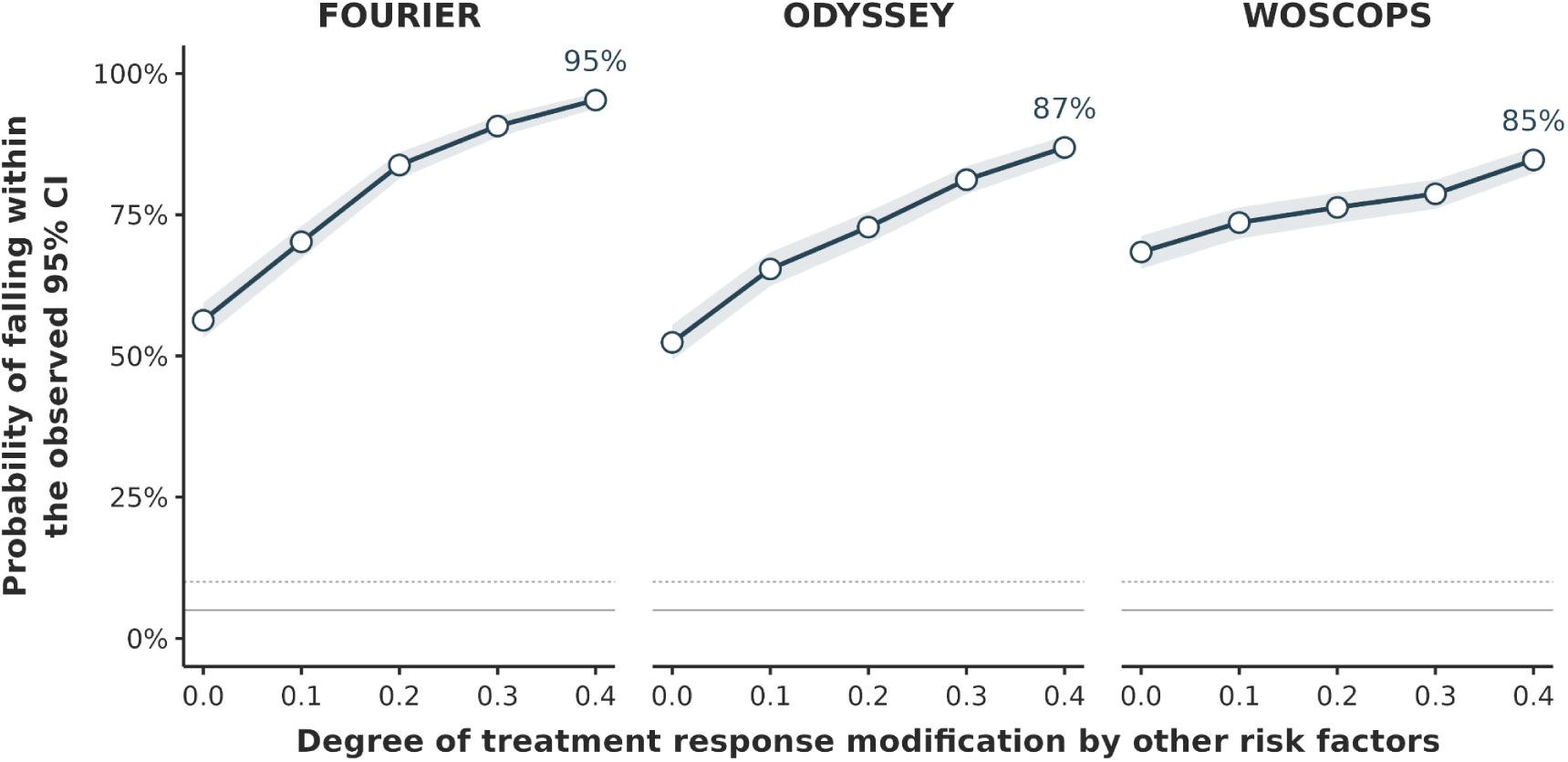
Compatibility probability for the published high-PRS versus lower-PRS treatment HR ratio as a function of treatment response modification by the other risk factor, across 1,000 simulations at the actual sample size of FOURIER (lek), ODYSSEY (centre), and WOSCOPS (right). Each point shows the percentage of simulations producing a high-versus-low PRS treatment HR ratio lying inside the 95% confidence intervals of the published ratios; shaded bands show simulation uncertainty. At zero treatment response modification, compatibility was 56% (95% CI 0.53 - 0.59) in FOURIER, 52% (95% CI 0.49 - 0.56) in ODYSSEY and 68% (95% CI 0.65 - 0.71) in WOSCOPS; at a treatment response modification of 0.40, this rose to 95% (95% CI 0.94 - 0.97), 87% (95% CI 0.85 - 0.89) and 85% (95% CI 0.82 - 0.87) respectively. The true PRS-by-treatment interaction is zero in all simulations.

### Observational analyses test predictions from the trial reanalysis simulations

The simulations reported in the previous section generate two testable predictions that can be evaluated in real-world clinical data. First, if the PRS-dependent treatment responses reported in the trial reanalyses were truly a consequence of trial eligibility criteria, as we hypothesised, then they should not be observed in a less selected study population. Second, we tested whether we could induce PRS-dependent treatment responses by applying trial-like eligibility criteria to the study population. We therefore evaluated these two testable predictions in UK Biobank (UKB) and Helix Research Network (HRN). We first addressed the original clinical question as directly as possible: whether a CAD PRS modifies the effect of statin prescription on acute coronary events. To improve power, we then examined whether an LDL PRS modifies LDL cholesterol response to statin prescription as a more proximal and quantitative outcome.

### Real-world observational analyses show limited evidence of modification of statin response by CAD PRS on coronary events

We first tested the most direct analogue of the retrospective trial analyses by asking whether CAD polygenic risk modified the effect of statin initiation on acute coronary events in UKB and HRN. Unlike FOURIER, ODYSSEY and WOSCOPS, these real-world clinical-care cohorts were not assembled using the restrictive combinations of cardiovascular history and LDL cholesterol that defined entry into the trials. We used IPTW within a target trial framework to estimate the effect of statin initiation across levels of CAD polygenic risk while balancing measured determinants of treatment initiation between initiators and non-initiators (**Methods**).

We found little evidence that CAD PRS modified the effect of statin prescription on acute coronary events (**Extended Data Fig. 3**). There was a nominally significant interaction in UKB for events within a 10-year follow-up window (95% CI OR 0.70–0.95, P = 0.01, N = 92,859), but this was not replicated in the larger HRN cohort. Despite its wide confidence intervals, the HRN analysis excluded PRS-stratified treatment response differences of the magnitude reported in the clinical trial reanalyses (95% CI OR 0.90 - 1.29, P = 0.40, N = 175,106). Taken together, these IPTW analyses did not recapitulate the large PRS-dependent treatment response reported in the trial populations, consistent with the first prediction from our simulations.

However, these analyses were less well-suited to testing the second prediction: that imposing trial-like eligibility criteria within a more representative population can generate apparent PRS-dependent treatment response. This is because there were relatively few clinical events and there was evidence of residual confounding because statin initiation was not randomized. To partially address both limitations, we next examined whether an LDL PRS modifies LDL cholesterol response to statin initiation. LDL cholesterol is the most pharmacologically proximal read-out of statin response and, as a continuous outcome, provides substantially greater statistical power. In addition, studies in clinical trials suggest that baseline LDL cholesterol level has a modest effect on relative post-treatment change in LDL cholesterol^21^. This provides a useful prior that the real-world data analysis in the UKB and HRN datasets should show limited effects of the LDL PRS on relative treatment response.

### Baseline LDL cholesterol stratification recreates trial-like collider bias in simulation

Before proceeding to the UKB and HRN analyses, we first established a ground truth reference for testing the second prediction by simulating routine-care statin prescription and LDL cholesterol response under a model in which the true effect of the LDL PRS on treatment response was fixed at zero (**Methods**). To ensure a realistic simulation setting, statin prescription was modelled to be non-random and depended strongly on baseline LDL cholesterol, as in routine clinical practice. We then compared the estimated PRS-by-treatment interactions in the full simulated population with estimates obtained after restricting the same population by baseline LDL cholesterol.

In the full simulated cohort, IPTW recovered the true null interaction (**Fig. 5**). By contrast, when the same cohort was stratified into tertiles of baseline LDL cholesterol, the estimated PRS-by-treatment interaction shifted away from zero, such that higher LDL PRS appeared to be associated with a blunted response to statin initiation. Because the true interaction was zero by construction, these observed subgroup effects were artefactual consequences of stratifying on baseline LDL cholesterol, analogous to trial selection. The simulation therefore provided a reference pattern for the empirical analyses, namely, little evidence of PRS-dependent treatment response in the unrestricted population, but apparent modification after imposing eligibility-like restrictions on a baseline measurement.

**Figure 5:**
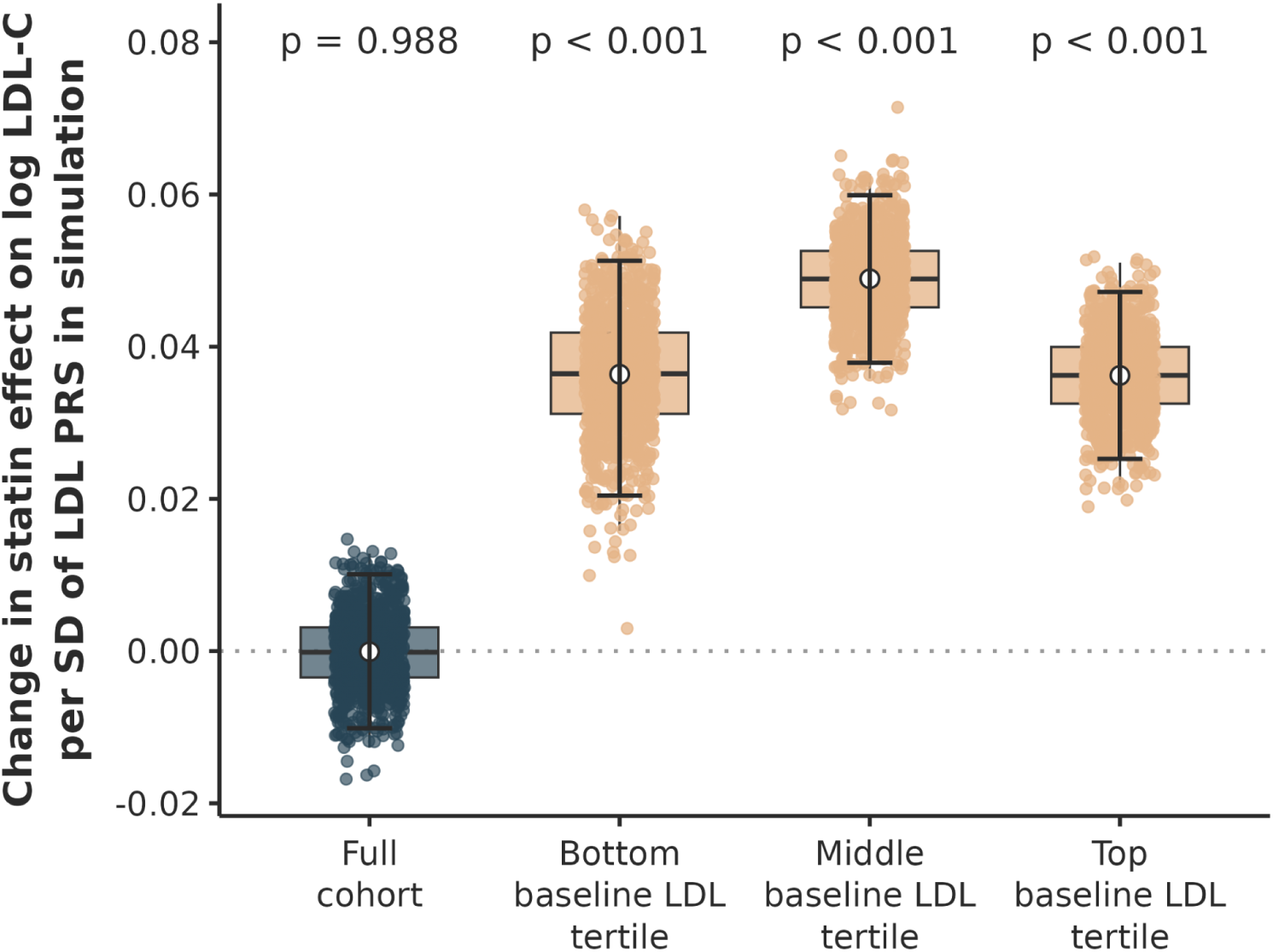
Estimated LDL PRS-by-statin initiation interaction coefficients from the observational cohort simulation, comparing full-cohort IPTW with IPTW applied within each tertile of baseline LDL cholesterol. Distributions reflect 1,000 Monte Carlo replications at the HRN sample size of 78,145. P-values correspond to the PRS-by-statin interaction estimates. The true PRS-by-treatment interaction is zero by construction. Positive values indicate that a higher LDL PRS is associated with a blunted LDL cholesterol response to statin initiation; negative values indicate greater response. In all analyses, the simulation was designed such that there was no PRS-by-treatment interaction.

### Imposing trial-like eligibility criteria in two real-world clinical-care cohorts induces apparent PRS-dependent treatment response

We next tested whether the pattern observed under known ground truth in the simulation was also evident in UKB (N = 28,299) and HRN (N = 78,145) for LDL cholesterol. As before, we emulated a target trial of statin initiation in UKB and HRN using IPTW to balance measured determinants of treatment initiation between initiators and non-initiators (**Methods**). In the primary IPTW analysis, statin initiation was associated with a 20–30% relative reduction in follow-up LDL cholesterol in both HRN and UKB (**Extended Data Fig. 4**), consistent with the lower range of LDL cholesterol reductions reported in randomized statin trials, potentially reflecting unfilled prescriptions in the real-world clinical-care data^22,23^. However, there was little evidence that LDL PRS modified this response. Estimates of the PRS-by-statin interaction were centred close to the null in both cohorts with tight confidence intervals.

We next stratified individuals by baseline LDL cholesterol tertiles as in the simulation, thereby creating trial-like subgroups analogous to studies that enrol participants only within restricted ranges of baseline LDL cholesterol level. In contrast to the full-cohort IPTW analysis, LDL-stratified analyses induced PRS-by-statin interactions in both cohorts (**Fig. 6; Extended Data Fig. 5**), indicating that a higher PRS corresponded to a blunted reduction LDL cholesterol response to statins across all tertiles.

**Figure 6:**
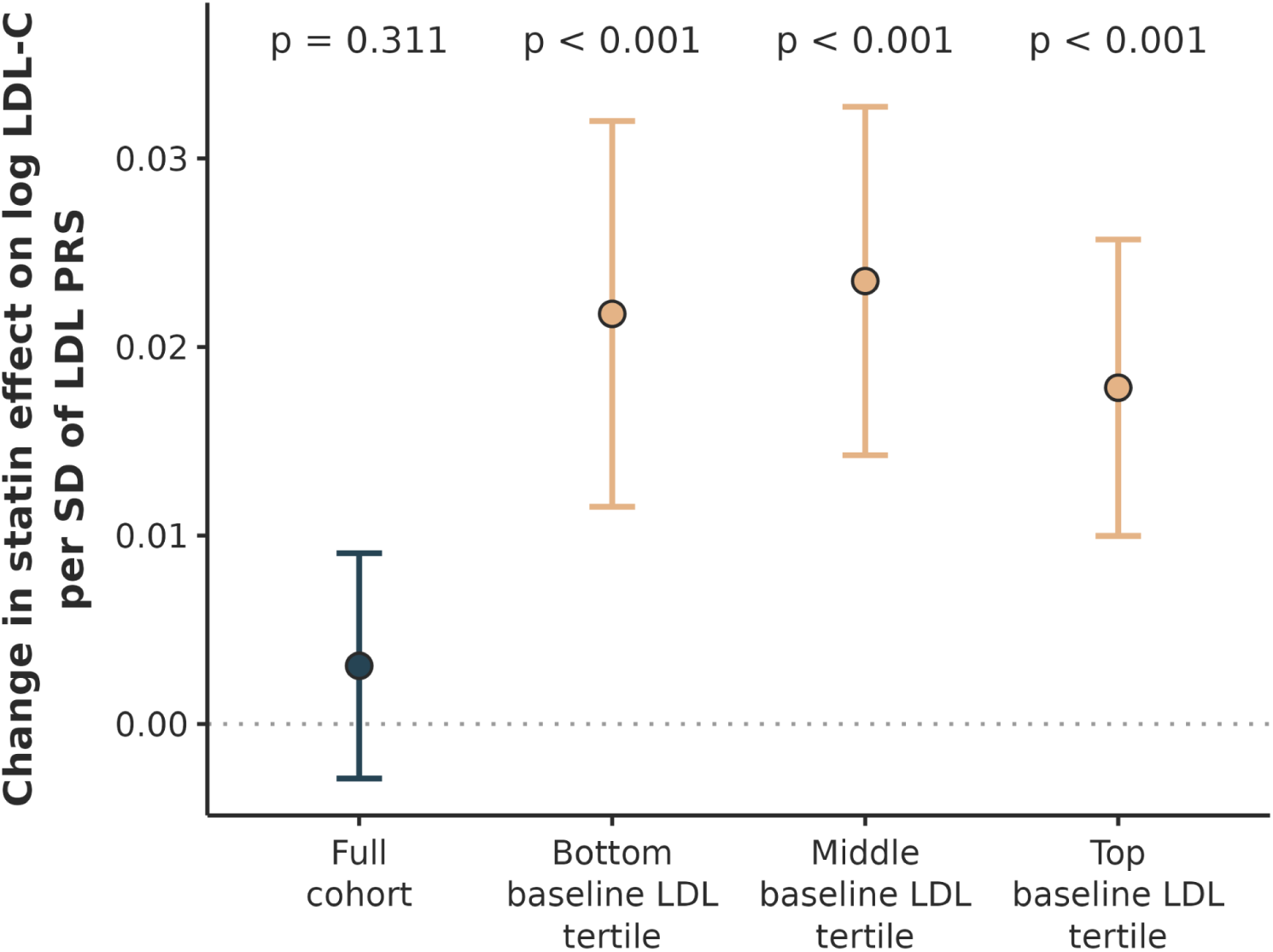
Interaction between statin initiation and LDL PRS on follow-up LDL cholesterol within the first, second and third tertiles of baseline LDL cholesterol. Each estimate is an inverse-variance weighted meta-analysis of cohort-specific estimates in UKB and HRN. P-values correspond to the PRS-by-statin interaction estimates. Within each cohort, IPTW is performed to balance covariates between initiators and non-initiators. Positive values of the interaction indicate that the LDL PRS is blunting LDL response to statins, while negative values indicate it is conferring greater response. Cross-cohort heterogeneity is low for the full cohort (I² = 23%, P = 0.25) and the top tertile (I² = 25%, P = 0.25), moderate for the middle tertile (I² = 64%, P = 0.10) and substantial for the bottom tertile (I² = 92%, P = 0.0006).

These results are consistent with the second testable prediction of our simulations. In two independent real-world clinical-care cohorts where the full data found little evidence of PRS-dependent statin response, imposing LDL-based eligibility criteria was sufficient to generate apparent PRS-by-treatment interactions with directions and magnitudes consistent with the simulation prediction (**Fig. 5**). Consistent results in both a simulation framework and empirical observational data provides important evidence that the biasing mechanisms described thus far can exist, and be detectable, in real-world data.

## Discussion

A central promise of personalized medicine is that response to treatment will depend, in part, on a patient’s genetic profile. Recent re-analyses of large-scale randomized controlled trials (RCTs) have reported substantial differences in response to lipid-lowering therapy according to polygenic risk for cardiovascular disease, resulting in highly favourable numbers needed to treat among individuals at high genetic risk. These findings have encouraged the use of PRSs—derived from GWAS of phenotypes such as CAD—to identify patients who may benefit most from treatment—hypotheses which are currently being tested in trials. This use is distinct from prognostic enrichment, in which PRSs are used to identify individuals at greater risk of experiencing trial outcomes and thereby increase trial efficiency; our findings concern the use of PRSs to predict differential treatment response. Here, we show that such patterns are also compatible with collider bias arising from trial eligibility, rather than requiring a direct biological effect of genetic risk on treatment response.

This is important because randomization may reasonably be assumed to ensure that differences in treatment response across patient characteristics reflect genuine biological effects of personalizing therapies to specific subsets of patients. However, randomization balances treatment arms only within the selected trial population; it does not remove collider associations among baseline characteristics induced by trial eligibility. In our simulations of FOURIER, ODYSSEY and WOSCOPS, eligibility induced associations between CAD PRS and other determinants of treatment response, causing the PRS to become a proxy for other response modifiers. Consequently, apparent PRS-dependent treatment effects arose despite randomized treatment assignment and a true PRS-by-treatment interaction of zero.

Within the collider mechanism we propose, CAD PRS can become a proxy for a true response modifier, making it tempting to conclude that PRS should generally predict greater treatment benefit. However, this proxy relationship is specific to the way participants were selected into FOURIER, ODYSSEY and WOSCOPS, where eligibility depended on downstream clinical characteristics such as cardiovascular disease and LDL cholesterol. The ongoing EE-PRS and PROACT2 trials select participants partly or primarily on the basis of high CAD PRS and therefore need not recreate the associations between PRS and other response modifiers induced in the retrospective trial populations. These trials may still benefit from prognostic enrichment, but the greater relative treatment response reported in the retrospective analyses should not be assumed to transport to populations selected using different eligibility criteria.

In real-world observational data, the problem takes a different form because observational studies are also prone to bias from confounding, which is generally absent in RCTs. In observational studies addressing the effect of CAD PRS on treatment response, baseline LDL cholesterol and cardiovascular history must be addressed as confounders of statin initiation, yet both variables lie downstream of genetic risk. Omitting them risks confounding, while conditioning on them can introduce collider bias. Our simulations confirmed this directly, since IPTW applied to the full cohort recovered estimates near the true null, whereas stratifying by baseline LDL cholesterol recreated the same artefactual pattern of PRS-by-treatment interaction observed in the trial simulations (**Fig. 5**). This provides a practical analytical approach for researchers using real-world clinical-care data to study PRS-dependent treatment response. Another common observational approach is to restrict analyses to drug initiators and estimate the association between the PRS and change in the outcome from before to after treatment; these designs can also be biased, including through a similar collider mechanism (**Supplementary Note 1**). Caution is still warranted, however, because IPTW remains vulnerable to residual confounding, as suggested by our CAD PRS analyses.

These predictions were borne out in both real-world cohorts as well. In the HRN and UKB, statin prescription produced a 20–30% reduction in LDL cholesterol, and target trial analyses found little evidence that the LDL PRS modified this response. Stratification by baseline LDL cholesterol generated apparent PRS-by-statin interactions in both datasets, consistent in direction and magnitude with the simulation. For coronary events, the CAD PRS showed some evidence of treatment modification in UKB, but this was not replicated in the larger HRN dataset, and the confidence intervals excluded the effect sizes reported in the trial reanalyses. Given the lower event counts and greater vulnerability to residual confounding in these observational analyses, this comparison should be interpreted cautiously. Therefore, the main contribution of this analysis has been to show that the large interaction effects reported in the retrospective trial reanalyses are not reproduced using an IPTW approach in routine care data. This interpretation is consistent with Cholesterol Treatment Trialists’ Collaboration meta-analyses showing that statin-induced relative reductions in major vascular events are broadly similar across baseline risk strata, whereas absolute benefit depends chiefly on baseline event risk^24,25^.

Several limitations of this work should be considered. Our simulations show that trial eligibility can induce collider bias sufficient to reproduce the findings from lipid-lowering trial reanalyses. However, they do not establish that it actually caused those original findings. The magnitude and direction of bias depend on simulation parameters that are difficult to verify empirically, especially the extent to which other risk factors collectively modify treatment response. The simulations are, therefore, best thought of as a plausible alternative explanation for the trial reanalyses’ findings. The observational analyses provided an independent empirical test of the collider bias mechanism, but the most precise results came from LDL PRS modifying the effect of statin initiation on relative change in LDL cholesterol. This is different from the trials, which explored the effect of CAD PRS modifying the reduction in coronary events. Our real-world clinical care analysis of CAD PRS was less precise and showed evidence consistent with residual confounding. Finally, and again, our findings are restricted to relative treatment effect modification; they do not undermine the use of PRSs for disease risk prediction or enriching trials for events, which may still enable smaller sample sizes and improve absolute risk reduction.

The broader implication of our findings is that personalized medicine approaches that rely on differential response to medicines due to genetic background should be carefully interpreted and may be subject to collider bias. Polygenic risk scores are determined before birth, and sit upstream of, and influence, a long series of clinical measurements, biomarkers, diagnoses and healthcare interactions, and are therefore especially vulnerable to bias from stratification and sample selection. While PRSs are clearly able to identify individuals at higher risk of disease, these findings suggest that caution should be taken if clinicians, trialists or patients expect that increased polygenic risk will translate to improved treatment response.

## Methods

### Simulation framework

#### Benchmarking against lipid-lowering trial results and eligibility criteria

Three published retrospective analyses of major lipid-lowering trials provide the quantitative benchmarks against which the simulations are evaluated. In the FOURIER reanalysis, the hazard ratio for major vascular events was 1.02, 0.85, and 0.75 in the lowest, intermediate, and highest quintiles of CAD PRS, respectively. In the ODYSSEY reanalysis, the corresponding hazard ratio for major adverse cardiovascular events was 0.87 among participants below the 90th percentile of CAD PRS and 0.63 above it. In the WOSCOPS reanalysis, the hazard ratio for incident coronary heart disease was 0.56 in the highest quintile of CAD PRS and 0.76 in all other participants. Dfobespite their directional consistency, these trials used structurally different eligibility criteria: FOURIER required established atherosclerotic cardiovascular disease together with LDL cholesterol ≥70 mg/dL or non-HDL cholesterol ≥100 mg/dL on high-intensity statin therapy; ODYSSEY required a recent acute coronary syndrome and LDL cholesterol ≥70 mg/dL, non-HDL cholesterol ≥100 mg/dL, or apolipoprotein B ≥80 mg/dL despite maximally tolerated statin therapy; and WOSCOPS enrolled men with LDL cholesterol ≥174 mg/dL but no history of myocardial infarction, selecting a primary-prevention population with the opposite cardiovascular history requirement. The fact that the same qualitative pattern of greater apparent benefit at higher PRS emerges across trials with opposite disease-history criteria motivates a structural explanation. We therefore asked whether a single data-generating model, in which only the eligibility parameters are changed between trials, could reproduce this pattern under a null treatment-by-PRS interaction.

#### Clinical trial simulation

In the simulated source population of *N* individuals, each individual was assigned a standardized CAD polygenic risk score *G* and a variable representing an aggregate of other risk factors, drawn independently from standard normal distributions:

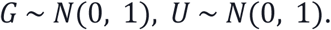

*U* represents a composite of other CAD risk factors that independently contribute to cardiovascular disease risk and to modification of treatment response. Within each randomised trial, *U* is mean-standardized.

Baseline log LDL cholesterol was generated as a linear function of *G* and *U*:

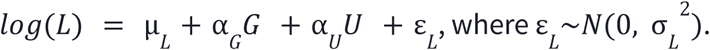

Cardiovascular disease liability *B* was generated as a linear function of *G*, *U* and *log*(*L*):

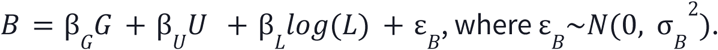

This formulation reflects that cardiovascular disease risk is multifactorial depending on genetics, LDL cholesterol and other clinical factors captured by the composite *U*. This is a deliberate modelling assumption: because *G* and *U* are independent in the source population but both contribute to trial eligibility, selecting a trial population on those variables can induce a negative correlation between them. Collider bias is model-dependent; under other ways in which these factors combine to determine trial eligibility, such as perfectly multiplicative/log-additive effects on the selected state, this induced correlation could be smaller, absent or even positive^26^.

Simulating both LDL cholesterol and cardiovascular disease liability enables us to mimic a simplified, but flexible, set of eligibility criteria overlapping those of FOURIER, ODYSSEY and WOSCOPS. Specifically, trial eligibility was imposed jointly on LDL cholesterol and cardiovascular disease liability. Let *q* _1−*p*_ denote the (1 − *p*)th quantile of *B* in the source population, where *p* naturally represents the trial-specific proportion of the source population meeting the cardiovascular disease history eligibility criterion. For FOURIER- and ODYSSEY-like trials, which required established or recent cardiovascular disease, eligibility was defined as:

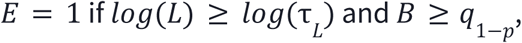

where

τ *_L_* denotes the trial-specific LDL threshold. For WOSCOPS, which excluded prior myocardial infarction:

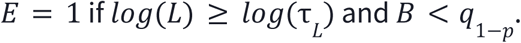

Treatment was assigned only after eligibility was determined and was randomized within the trial-eligible population:

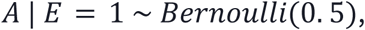

ensuring that treatment assignment *A* is independent of all pre-randomization variables given trial eligibility.

The primary endpoint *Y* was generated as a binary event indicator using a Cox proportional hazard model:

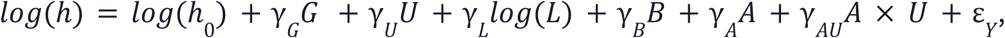

where 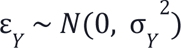 and event times were drawn as *T* ∼ *Exponential*(ℎ) and the actual event *Y* = 1 if *T* ≤ 1. The baseline cumulative hazard ℎ is not a fixed intercept, rather, it is calibrated numerically within each simulation run via root-finding to match the trial-specific placebo event rate. The term γ*_AU_* allows the log treatment hazard ratio to shift linearly with the other risk factor *U*, representing clinically plausible heterogeneity in the treatment benefit driven by non-genetic factors. Crucially, there is no *G* × *A* interaction term in this model, such that the true treatment-by-PRS interaction is zero by construction, so any estimated interaction in the simulated trial data must arise entirely from statistical bias.

It is important to note that *U* has twin properties: it must both contribute to trial eligibility and modify treatment response. The first property creates a collider-induced association with the PRS *G* within the trial population and the second property induces a spurious *G* × *A* interaction. If *U* affects trial eligibility but does not modify treatment response, it may become associated with *G* within the trial population, but this is insufficient to generate a spurious interaction.

Each trial simulation was run 1,000 times at the corresponding trial sample size, sampling a new source population of 300,000-1,500,000 individuals at each iteration before applying trial eligibility. PRS-stratified hazard ratio estimates, structured to match the PRS subgroups in the corresponding retrospective trial analyses, were obtained within each simulated trial using a Cox proportional hazards estimator, analogous to the published analyses. Results are summarised as means and 2.5-97.5th percentile intervals across simulation runs. Compatibility with the published PRS-stratified findings was assessed as the proportion of simulation runs in which the ratio of high-PRS to low-PRS treatment hazard ratios lay inside the 95% confidence interval of the published values (**Extended Data Fig. 2**). Formally, the compatibility probability 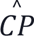 is calculated as:

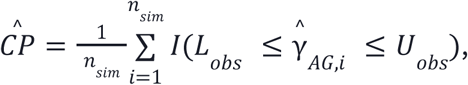

where 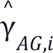 is the estimated high-PRS versus low-PRS HR ratio in each simulation run *i* and *L _obs_* and *U _obs_* are respectively the lower and upper bounds of the 95% confidence interval of the corresponding published HR ratio from the trial reanalysis, and *n _sim_* = 1, 000.

Parameter values were anchored to published trial characteristics, external epidemiological estimates or observed quantities wherever possible. For parameters that could not be directly estimated, we specified clinically plausible anchor values and ranges and evaluated their influence in sensitivity analyses (**Tables 1–2**).

#### Sensitivity of non-genetic treatment response modification

The key parameter in our simulation is γ : the extent to which the non-genetic CAD risk factor modifies response to treatment. We therefore varied this parameter from 0 to 0.40 and, at each value, calculated the proportion of simulations in which the high-versus-lower PRS treatment HR ratio fell within the 95% confidence interval of the corresponding published trial estimate (**Fig. 4**).

The range was chosen to encompass clinically plausible heterogeneity in the response to lipid-lowering therapy. For example, ODYSSEY reported that alirocumab’s hazard ratio for total cardiovascular events varied from 0.95 in the lowest baseline Lp(a) quartile to 0.75 in the highest quartile, corresponding to a treatment-modification parameter of approximately 0.18 in our model^27^. This parameter is on the log-hazard scale. Thus, a value of 0.18 corresponds in magnitude to an approximately 1.20-fold increase in the treatment hazard ratio per standard deviation increase in non-genetic risk, while a value of 0.40 corresponds to an approximately 1.49-fold increase.

Since the other risk factor represents a composite of many factors that may modify effective clinical benefit, including lifestyle factors, comorbidities, adherence and background lipid-lowering response, we selected a maximum value of 0.40 as a substantial but reasonable upper value for the sensitivity analysis. At this value, treatment hazard ratios were approximately 0.61 and 1.05 at the 25th and 75th percentiles of non-genetic risk.

#### Observational study simulation

The observational simulation shared much of the structure of the clinical trial simulation, but adapted it in three important ways. First, it removed randomised treatment assignment, and instead allowed treatment to depend on LDL cholesterol and, more weakly, the LDL PRS. Second, it did not have explicit eligibility criteria, instead using the source population directly, as in a representative population biobank or EHR. Third, it changed from a binary event endpoint to a continuous endpoint mimicking that of follow-up LDL cholesterol, to align it with the real data analyses.

The LDL PRS *G*, other risk factor *U* and log LDL cholesterol *log*(*L*) were generated via the same structure as the clinical trial simulation. It differed in that treatment initiation followed a logistic model depending on log LDL cholesterol and the LDL PRS:

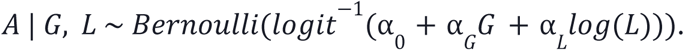

The other risk factor *U* did not directly influence *A* given *log*(*L*) and *G*, so that IPTW would be sufficient to remove confounding of treatment initiation. Follow-up LDL cholesterol was generated as:

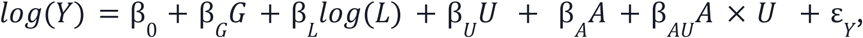

where β *_AU_* allows the other risk factor *U* to modify LDL response to treatment.

The simulation cohort was analysed using IPTW with stabilized weights *w* = *P*(*A*)/*P*(*A* | *G*, *log*(*L*)), constructed from true propensity scores. The simulation was run over 1,000 Monte Carlo replications at the HRN sample size of 78,145. To further evaluate the impact of baseline LDL stratification, IPTW was applied within each tertile of baseline LDL. This design mirrors the trial eligibility mechanism by restricting the analysis to a narrower range of a baseline clinical measurement (**Fig. 5**). See **Table 3** for a list of parameter values and their justification.

**Table 3:**
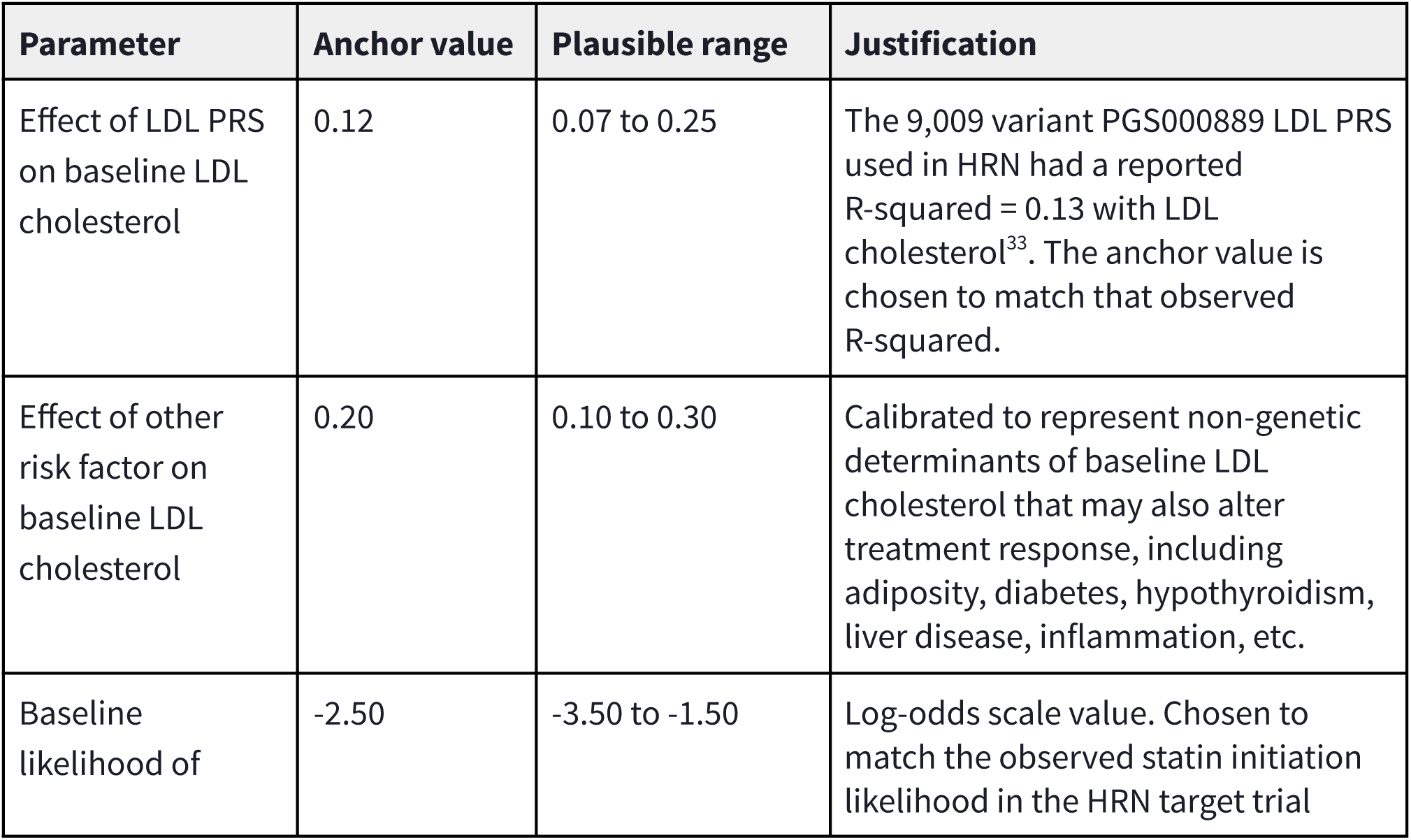
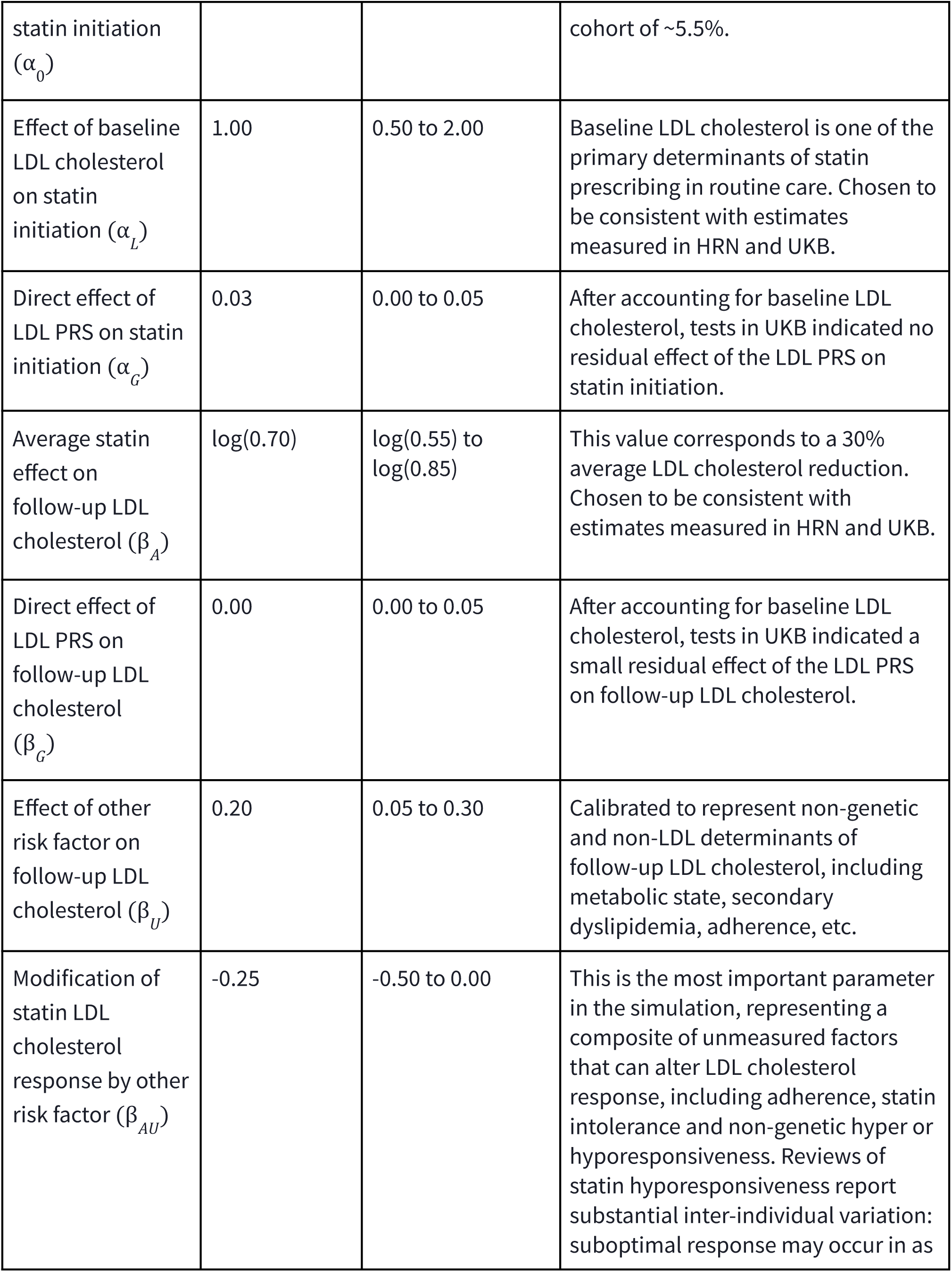
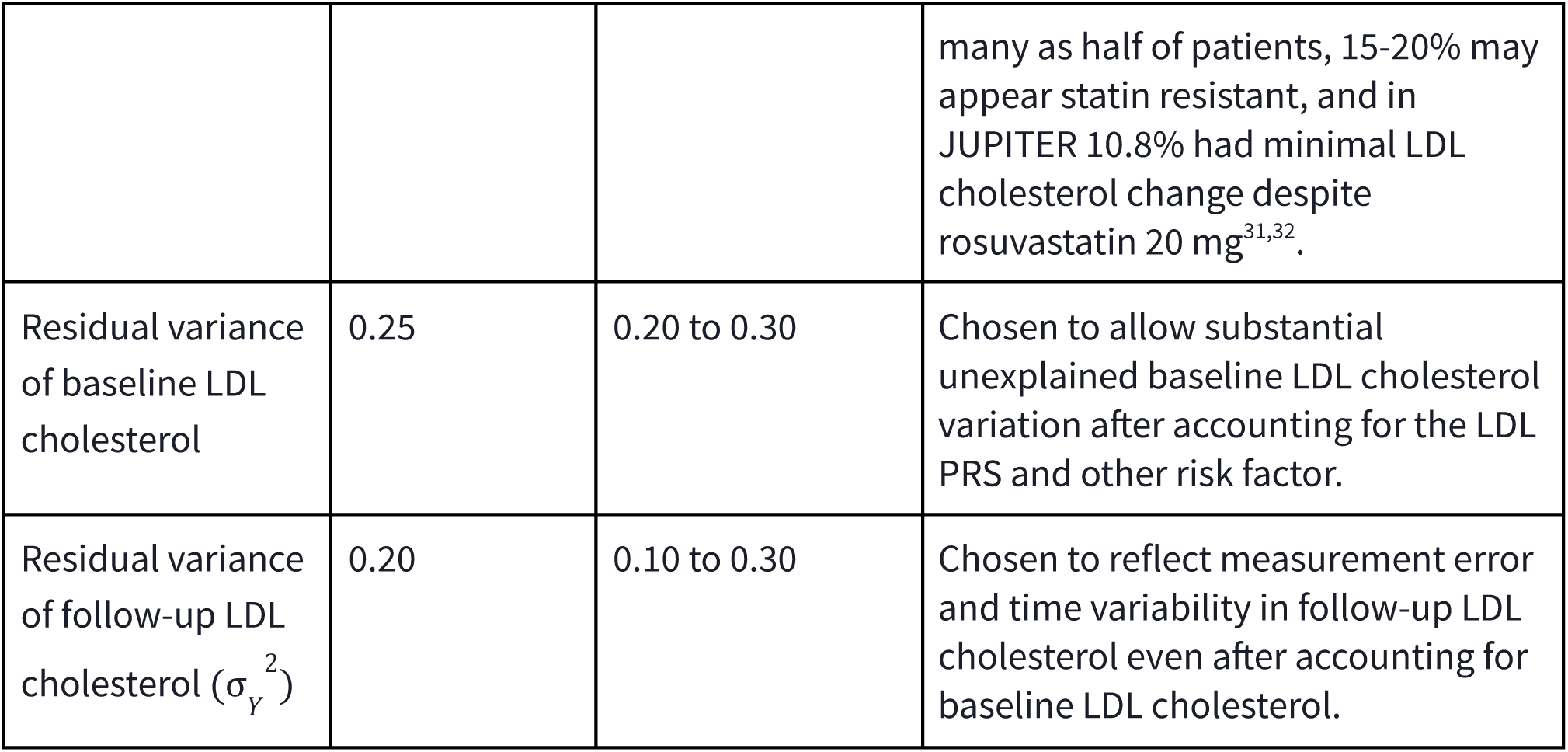
Simulation parameters for the observational biobank/EHR simulation.

### Clinical trial and observational study simulation parameters

#### Directed acyclic graphs showing bias in observational studies of PRS modification of treatment response

**Figure 7:**
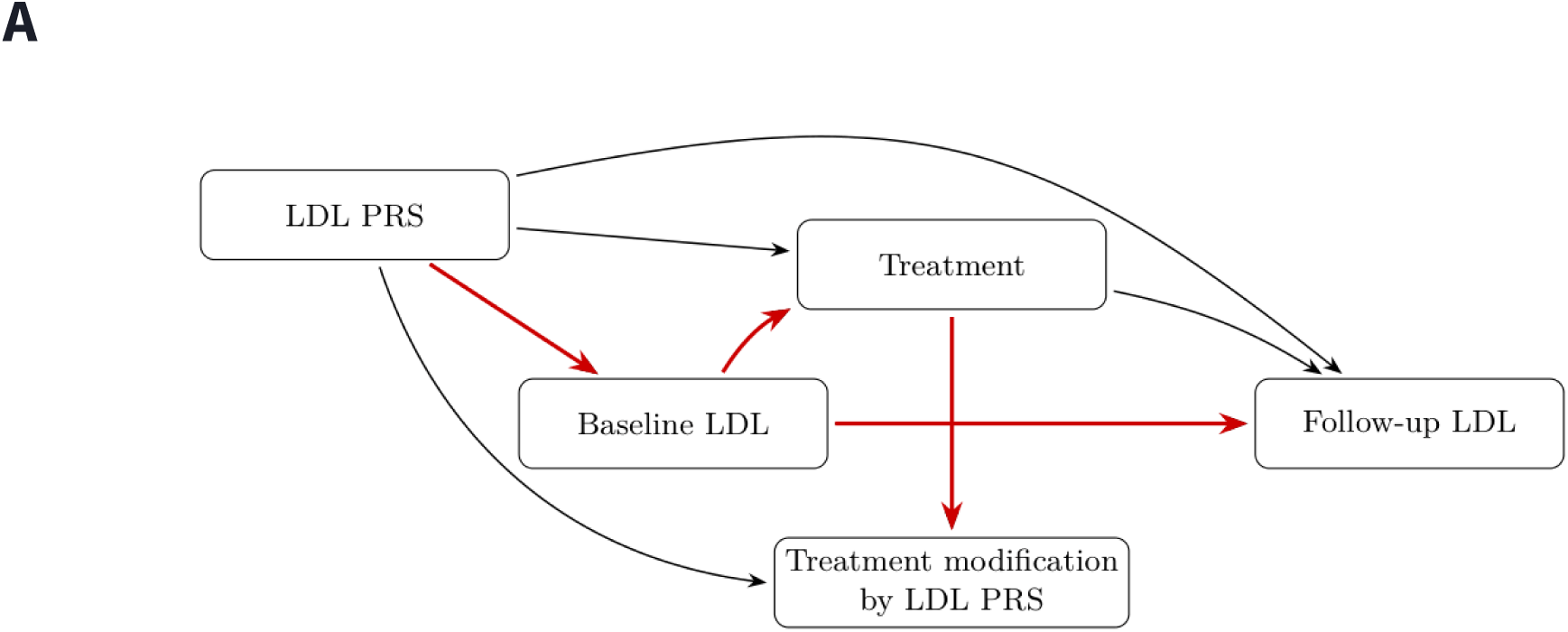
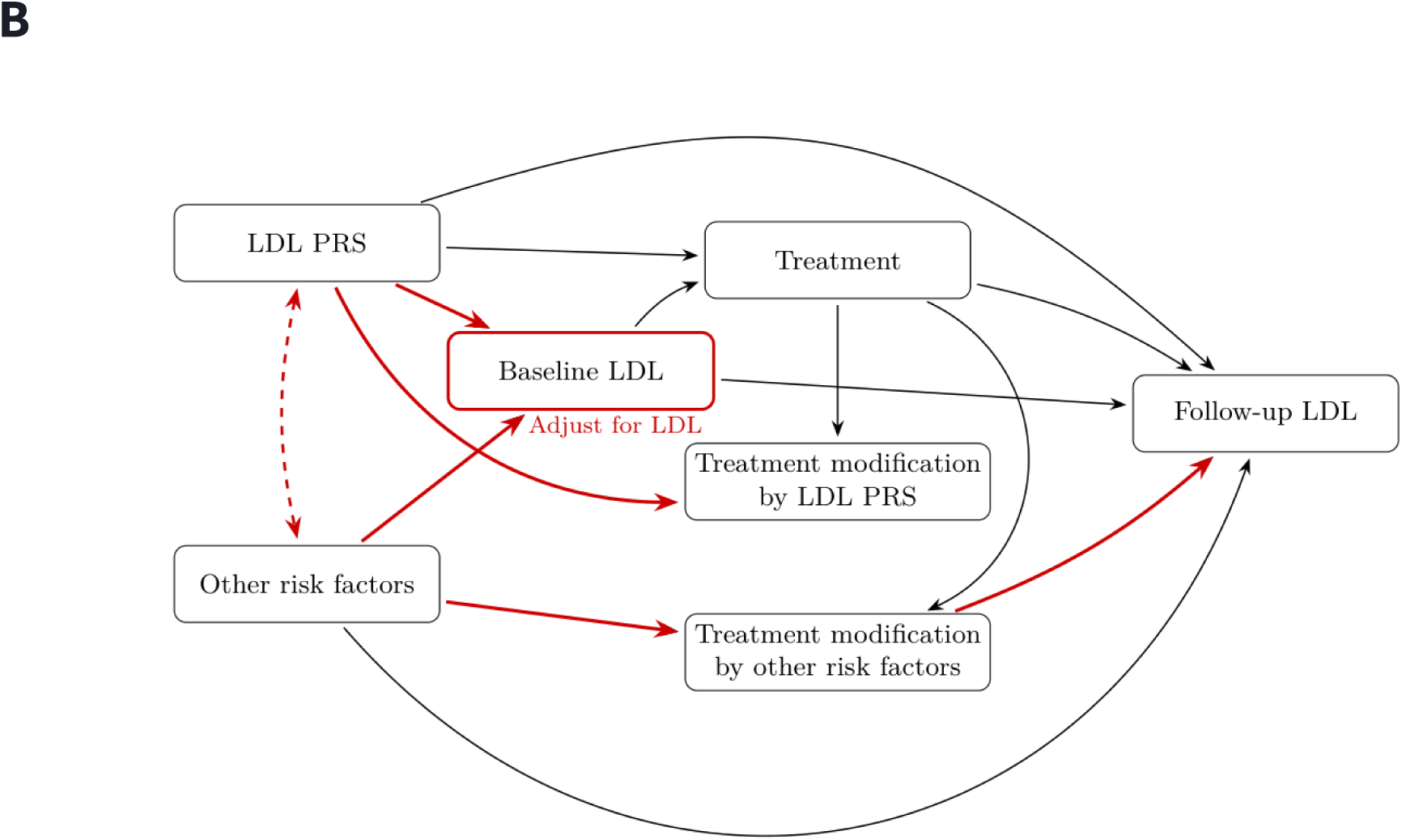
**A**, In observational cohorts, treatment is not randomized. Baseline LDL cholesterol may therefore confound estimates of LDL cholesterol PRS-by-treatment interaction because it is influenced by the PRS, affects treatment initiation and predicts follow-up LDL cholesterol. **B**, Adjusting for, stratifying on or restricting by baseline LDL cholesterol can instead introduce collider bias, because baseline LDL cholesterol is influenced by both the PRS and unobserved determinants of lipid metabolism, comorbidity or treatment response. Conditioning on baseline LDL cholesterol can therefore induce an association between the PRS and unobserved treatment-response modifiers.

### Real data analysis in Helix Research Network and UK Biobank

#### A target trial emulation framework for statin response in Helix Research Network and UK Biobank

Our simulations demonstrated that inverse probability of treatment weighting (IPTW) within a marginal structural model (MSM) can recover an unbiased estimate of the interaction between statin initiation and genetic risk, even when other plausible approaches are biased. However, in practice, appropriate statistical modelling must be complemented by robust study design. If eligibility, treatment assignment and follow-up are temporally misaligned or poorly defined, an MSM will also produce biased estimates. In large healthcare databases such as Helix Research Network (HRN), with medical and pharmacy EHR as well as claims data obtained through Komodo Health, and UK Biobank (UKB) primary care records, study designs that do not consider monitoring, ascertainment, and measurement norms for such datasets can lead to spurious findings. In this section, we provide a high-level overview of the study design applied in both HRN and UKB.

Target trial emulation is a conceptual approach to observational study design that addresses these problems. As articulated by Hernán and colleagues and illustrated in pharmacoepidemiologic applications such as the re-analysis of statins and cancer risk by Dickerman et al., the approach begins by specifying the protocol of the hypothetical randomized trial that would answer the causal question of interest: who is eligible, what treatment strategies are compared, when treatment is assigned, when follow-up begins and how outcomes are defined^16,17^. The observational analysis is then structured to mirror this protocol as closely as possible.

The need for this structure is particularly acute when studying gene-treatment interactions. A polygenic risk score is determined at conception, preceding all clinical measurements. In contrast, statin initiation occurs decades later, typically after multiple clinical encounters and laboratory assessments, and is strongly influenced by those measurements. We are therefore interacting an exposure that precedes an individual’s entire clinical history with a treatment decision that sits downstream of many intermediate clinical measurements. This complex time ordering requires a clearly defined eligibility window, treatment assignment period and start of follow-up to ensure that the resulting estimates have a clear causal interpretation.

In the present study, we are asking whether clinical response to statin initiation on both LDL cholesterol and incident CAD differs according to background polygenic risk. This is inherently a causal question. We therefore aim to specify and emulate a target trial in which individuals become eligible for inclusion if they satisfy (1) having a clinical LDL cholesterol measurement (corresponding to *L*), (2) a 12 (UKB) or 24 (HRN)-month wash-out period during which no statins were prescribed and (3) actively engaging with a primary care provider over this period, as evidenced by recorded encounters or measurements. The third criterion is to ensure that a lack of statin prescriptions does not simply reflect an absence of engagement with a given healthcare provider.

Among eligible individuals, we define a 12-month period following baseline during which treatment assignment is determined. Individuals receiving a statin prescription in this window are classified as initiators (corresponding to *A* = 1), and those who do not as non-initiators (corresponding to *A* = 0). Two aspects of this design are important. Firstly, we do not restrict to initiators, allowing statin initiation to be modelled as a treatment assignment mechanism within the MSM, depending on baseline LDL cholesterol, age, sex, prior CAD and other characteristics. Secondly, we do not assume adherence to a statin prescription. In this target trial, *A* represents assignment to treatment; accordingly, our estimates correspond to an intention-to-treat (ITT) design.

Finally, after the 12 month treatment assignment period, we identify the subset of individuals who have either a follow-up LDL cholesterol measurement within the subsequent 12 month period, or a CAD event within the subsequent 10 year period.

|  | UKB | HRN |
| --- | --- | --- |
| *Y*: LDL cholesterol (follow-up) | ReadV2 and CTV3/ReadV3 code “44P6”. Restricted to 1-8 mmol/L. | EHR measurements with concept IDs of 3009966, 3028288 or 3028437. Restricted to 1-8 mmol/L. |
| *Y*: AMI | Primary hospitalisation or death due to ICD-10 I21-I24 within 10 years of the end of the statin initiation period. | Primary hospitalization due to SNOMED equivalents of ICD-10 I21-I24 (see supplement) |
| *A*: Statin initiation | BNF code starting with 02.12.1 or 2.12.1, or string matching on “atorvastatin\|simvastatin\|rosuvastatin\|pravastatin\|fluvastatin\|lovastatin\|pitavastatin”. | 25,456 RxNorm/RxNorm Extension concepts covering all dose forms, branded products, and combination products for: atorvastatin, simvastatin, pravastatin, rosuvastatin, lovastatin, fluvastatin, pitavastatin, cerivastatin. Combinations with ezetimibe, niacin, amlodipine, and others are also included. Patients were required to have two documentations of statin initiation (either both claims and EHR within 30 days, or twice in only claims or EHR at least 90 days apart). |
| *L*: LDL cholesterol (baseline) | ReadV2 and CTV3/ReadV3 code 44P6. Restricted to 1-8 mmol/L. | EHR measurements with concept IDs of 3009966, 3028288 or 3028437. Restricted to 1-8 mmol/L. |
| *G*: PRS | UK Biobank genetic score data-field 26250 (LDL cholesterol PRS), 26227 (CAD PRS), 26223 | PGS000889 (LDL PRS), PGS003446 (CAD PRS) |
|  | (CVD PRS) |  |
| Other covariates | Pre-existing CAD prior to baseline LDL cholesterol measurement (ICD-10 I21-I25); prior statin prescription before wash-out period; number of primary care visits and prescriptions in the 12 months prior to baseline LDL cholesterol measurement. | Pre-existing CAD prior to baseline LDL cholesterol measurement (see SNOMED list in supplement); prior statin prescription before wash-out period; number of prescriptions in the 12 months prior to baseline LDL cholesterol measurement. |

#### Propensity score weighting in Helix Research Network and UK Biobank

Within the target trial framework, propensity scores were used to construct inverse probability of treatment weights that balance observed covariates between statin initiators and non-initiators, approximating randomized treatment assignment. Propensity scores estimate the probability of statin initiation given observed baseline covariates, and were estimated using cross-validated LASSO logistic regression (R package glmnet), with the penalty parameter selected by ten-fold cross-validation minimizing binomial deviance at the cross-validated minimum (λ *min*). In the primary analysis, covariates entering the propensity model comprised log-transformed baseline LDL-C, age at the landmark date, prior healthcare utilization in the 365 days preceding the landmark (days with a dispensed prescription and days with a clinical contact), sex, prior statin use (none; 0–2 years; 2–5 years; >5 years before landmark), prior coronary artery disease, and the LDL-C and CAD polygenic risk scores. In the assessment-centre sensitivity analysis, the covariate set was expanded to additionally include log-transformed triglycerides, C-reactive protein, HbA1c, ALT and HDL-C; apolipoprotein B; uric acid; systolic and diastolic blood pressure; BMI; body fat percentage; Townsend deprivation index; smoking status; alcohol use; chest pain; diabetes; and educational attainment.

Stabilized inverse probability of treatment weights were constructed as *w* = *P*(*A*)/*P*(*A* | *X*), where the numerator is the marginal treatment prevalence estimated as the sample proportion of initiators and the denominator is the individual predicted initiation probability from the LASSO model. Weights were symmetrically truncated at the 5th and 95th percentiles to limit the influence of extreme values; the resulting distributions were narrow in both analyses (interquartile range approximately 0.88–1.05 in the primary analysis and 0.95–1.02 in the sensitivity analysis), indicating good covariate overlap between treatment groups. Balance was assessed graphically using a calibration plot of observed versus predicted initiation rates across twenty equal-frequency bins of the propensity score, and overlapping density histograms of propensity scores stratified by treatment status (**Fig. 8**).

**Figure 8:**
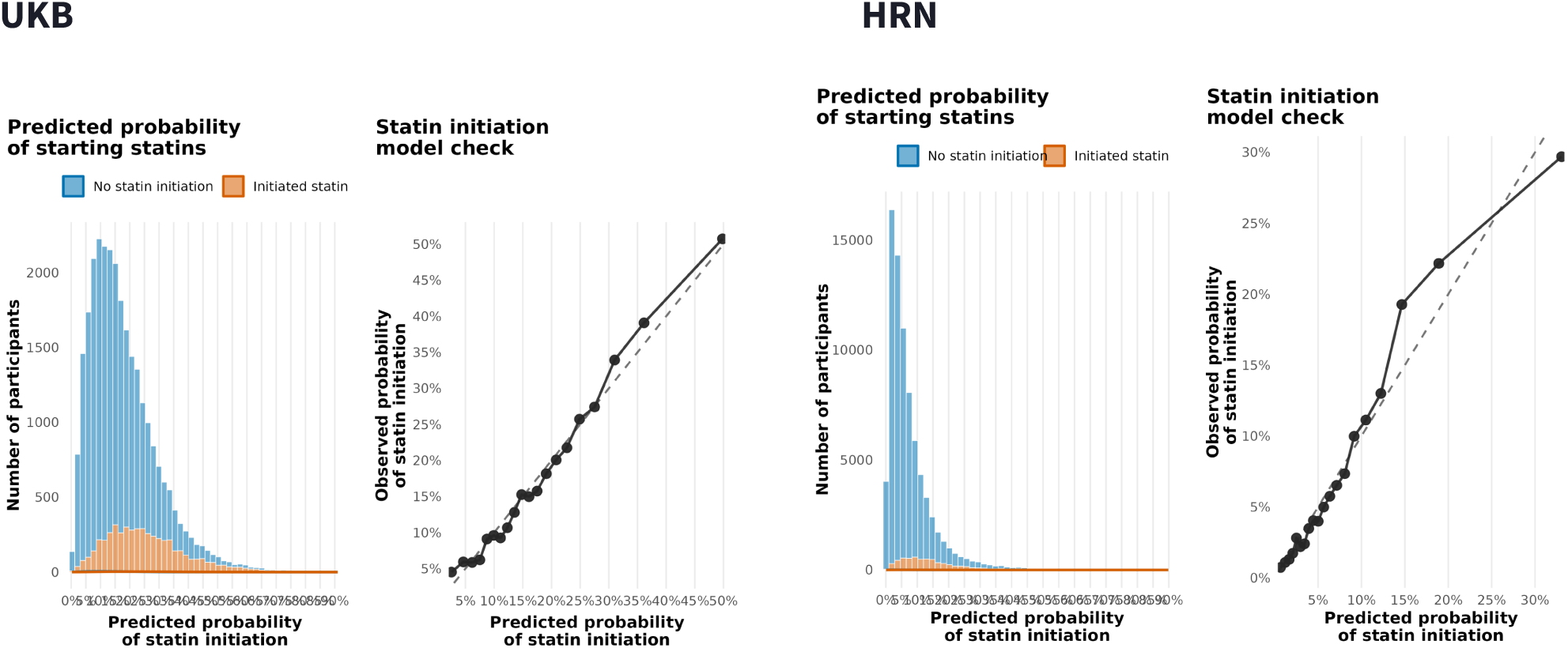
Propensity score diagnostics for the primary inverse probability of treatment weighting analysis in UKB primary care (lek) and HRN (right). Lek panels show calibration plots of observed versus predicted statin initiation rates; points lying close to the diagonal indicate that the LASSO propensity model is well-calibrated across the range of predicted values. Right panels show overlapping density histograms of propensity scores stratified by statin initiators and non-initiators; substantial overlap between distributions indicates that both groups are represented across the range of propensity scores, supporting the positivity assumption.

Participants without a follow-up LDL-C measurement were excluded by complete-case restriction; inverse probability of censoring weights were not applied. All analyses were restricted to participants of genetically confirmed European ancestry and were not pooled across other genetic similarity groups. Weighted outcome models for LDL-C used ordinary least squares regression on the log scale; those for incident coronary events used a quasi-binomial generalized linear model. Standard errors were derived from the model-based variance in both cases. The final acute coronary event analysis included 92,859 participants in UKB and 175,106 in HRN, whereas the LDL cholesterol response analysis included 28,299 participants in UKB and 78,145 in HRN.

## Data availability

UK Biobank data are available to eligible researchers following application to and approval by UK Biobank. The data that support the findings of this study were generated through the Helix Research Network (HRN), a multi-center research program governed by a Joint Steering Committee and Science and Research Subcommittee. The HRN Database is not publicly available due to participant privacy protections, institutional data governance requirements, and ethical restrictions. Qualified researchers affiliated with HRN Member Sites or approved third-party institutions may request access to de-identified data through the HRN’s established governance process, which includes submission of a research proposal, subcommittee review, completion of human subjects research training, and execution of a data use agreement. Inquiries regarding data access may be directed to. All simulated datasets and analyses can be reproduced using the code and parameters available in the public GitHub repository.

## Code Availability

Code used to reproduce the simulation analyses and UK Biobank routine-care analyses is publicly available in the following GitHub repository: https://github.com/matt-tudball/prs-drug-response.

## Acknowledgements

This research was conducted using data from the Helix Research Network under approved application 20250641. This research was possible because of the participants who have contributed to the Network through participating institutions: DNA Answers (St. Luke’s University Health Network, Pennsylvania, Christopher N. Chapman), DNA Discovery (TriHealth, Ohio, James F. Maher), DNA Insights (Parkview Health, Indiana and Ohio, Jamie L. Renbarger), GeneConnect (Cone Health, North Carolina, Chad Haldeman-Englert), the Genetic Insights Project (Nebraska Medicine, Douglas Stoller), GenoWell (Rochester Regional Health, New York, Pradyumna Phatak), the Healthy Nevada Project (Renown Health, Nevada, Joseph J. Grzymski), ImagineYou (Sanford Health, Upper Midwest, Steven F. Powell), In Our DNA SC (Medical University of South Carolina, Daniel P. Judge), myGenetics (HealthPartners, Minnesota, Douglas A. Olson), Ohio State Genomic Health (The Ohio State University Wexner Medical Center, Amy C. Sturm), PreciselyYou (WakeMed, North Carolina, William H. Lagarde), The Gene Health Project (WellSpan Health, Pennsylvania, C. Anwar A. Chahal), OurGenes (University Health Network, Ontario, Canada, Raymond H. Kim) and genoME (Memorial Hermann Health System, Texas, James McCarthy).

## Author contributions

M.J.T., J.B.R., E.T.C. and M.V.H. conceptualized the study and contributed to study design. M.J.T. designed the simulation framework. M.J.T. and E.T.C. performed the data analyses. M.J.T. drafted the manuscript. J.B.R., E.T.C. and M.V.H. contributed to manuscript development and revision. G.D.S. contributed to study design and critically reviewed the manuscript. All authors reviewed and approved the final manuscript.

## Ethics

The Helix cohorts were reviewed by Salus IRB (Reliance on Salus for all sites) and approved (approval number 21143), the WCG IRB (Western Institutional Review Board, WIRB Copernicus Group) and approved (approval number 20224919), the MUSC Institutional Review Board for Human Research and approved (approval number Pro00129083), and the University of Nevada, Reno Institutional Review Board and approved (approval number 7701703417). All participants gave their informed, written consent before participation. All data used for research were de-identified. The UK Biobank analyses reported here were conducted under application number 27449. The HRN analyses were conducted under approved HRN application 20250641. Simulation analyses involved no human participants and required no additional ethics approval.

## Extended Data

**Extended Data Figure 1:**
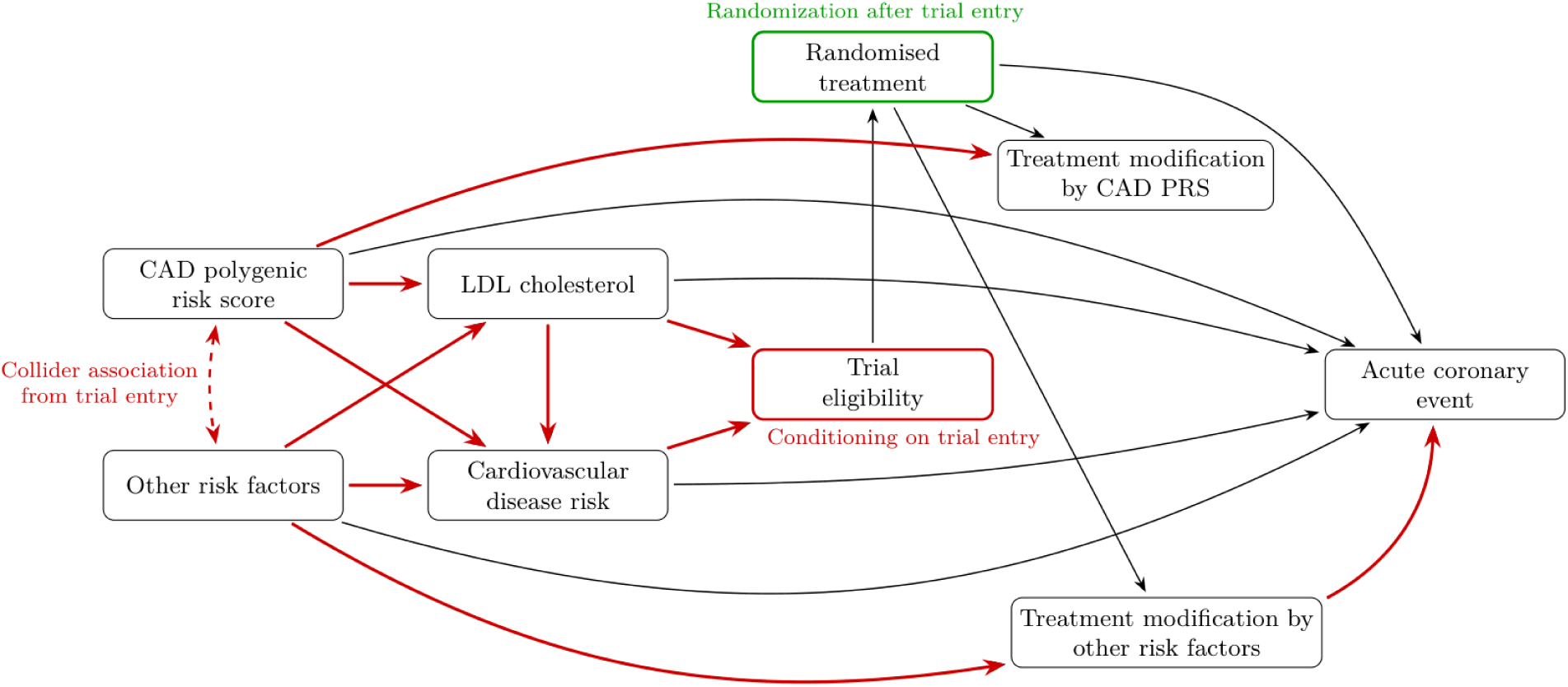
Directed acyclic graph showing the data-generating process used for the FOURIER-, ODYSSEY- and WOSCOPS-like simulations. In the source population, CAD polygenic risk and a non-genetic CAD risk factor were pre-specified to be independent from each other. Both contribute to LDL cholesterol levels and to cardiovascular disease risk. Trial eligibility is then imposed by restricting to individuals satisfying trial-specific combinations of LDL cholesterol and other inclusion criteria: established or recent cardiovascular disease with elevated LDL cholesterol for FOURIER- and ODYSSEY-like trials, and high LDL cholesterol without prior clinical disease for the WOSCOPS-like trial. Conditioning on eligibility creates an induced association between CAD PRS and non-genetic risk, shown by the dashed red arrow. Treatment is randomized aker trial entry, and therefore has no incoming arrows from baseline variables. The outcome model allows treatment benefit to vary with non-genetic risk but contains no true treatment-by-PRS interaction. Therefore, any apparent PRS-related treatment benefit estimated within the simulated trial arises from eligibility-induced selection rather than biological modification of treatment response by CAD polygenic risk. Red arrows indicate pathways contributing to the collider bias mechanism; the dashed red arrow specifically represents the induced association between CAD PRS and non-genetic risk aker conditioning on trial eligibility; the green node indicates randomized treatment assignment aker eligibility.

**Extended Data Figure 2:**
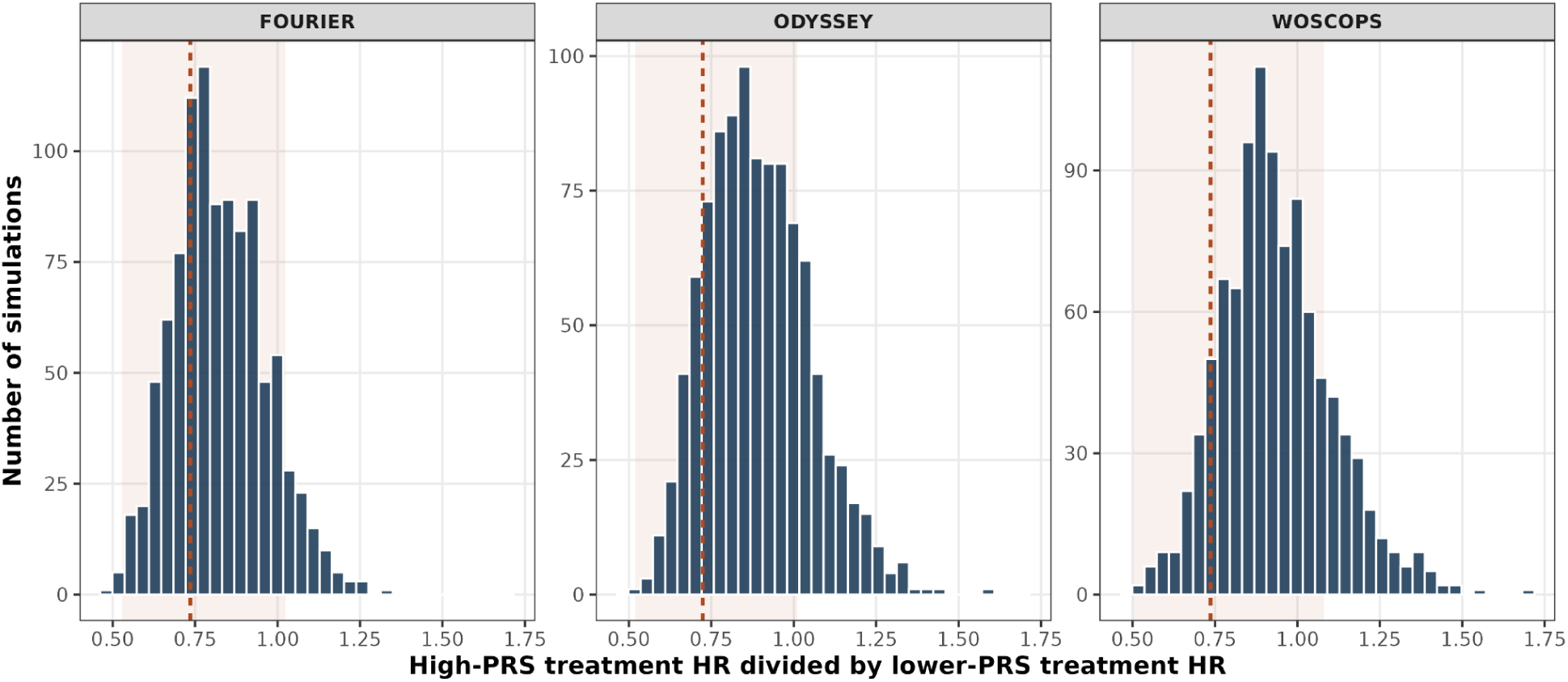
Distribution of the ratio of the high-PRS treatment HR versus lower-PRS treatment HR across 1,000 simulations, where the PRS-by-treatment effect is pre-specified to be zero. Values on the x-axis below 1 indicate apparently greater relative treatment benefit in the high-PRS subgroup. Each simulation run uses the actual sample size of FOURIER (lek), ODYSSEY (centre), and WOSCOPS (right). Dashed vertical lines indicate the published subgroup HR ratio for each trial and shaded areas mark the corresponding 95% confidence interval. The proportion of simulations lying inside the 95% confidence intervals of the published HR ratios was 91% in FOURIER, 81% in ODYSSEY and 79% in WOSCOPS.

**Extended Data Figure 3:**
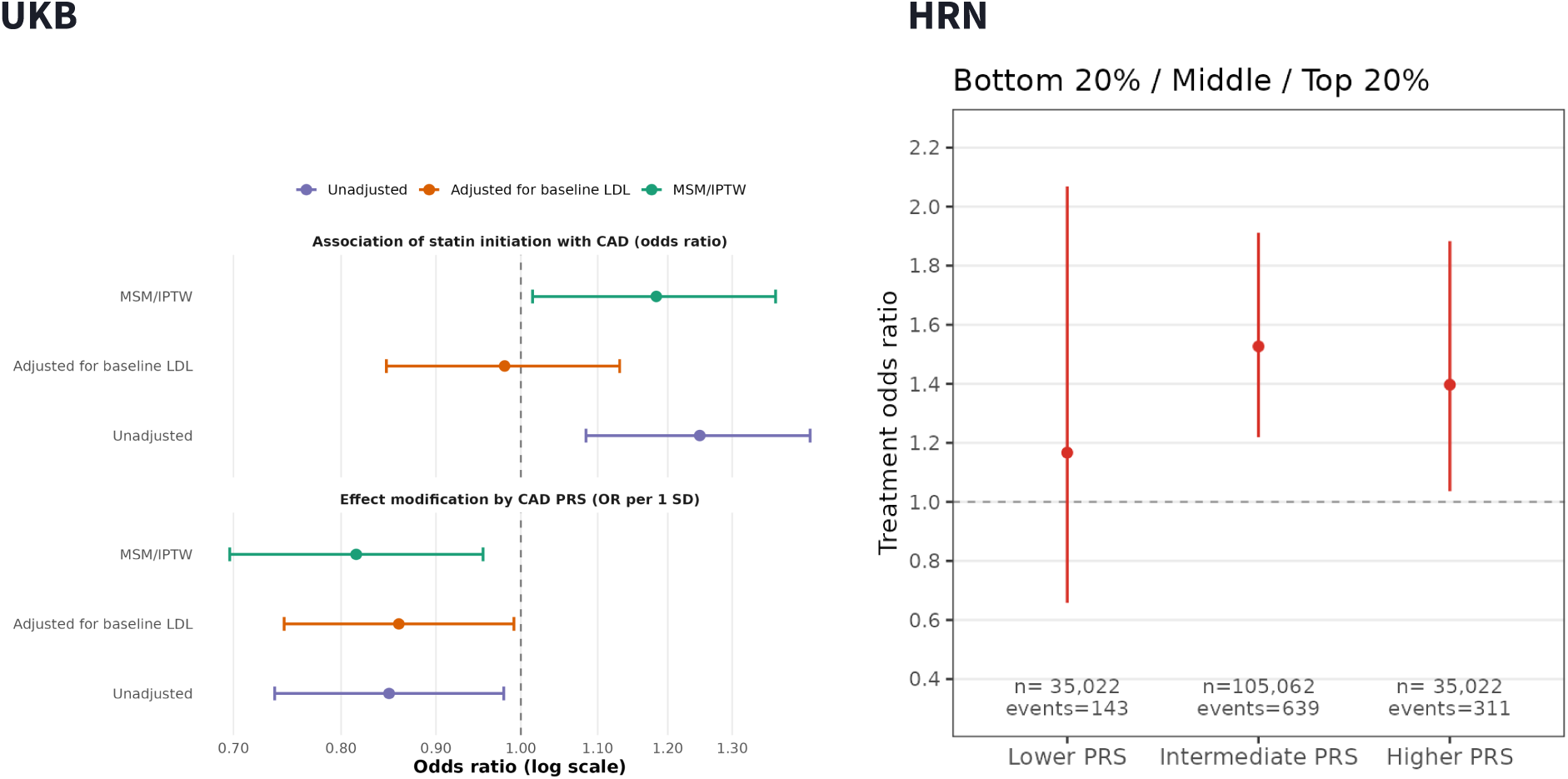
Secondary analysis of UKB primary care (n = 92,859) and HRN electronic health records (n = 175,106), testing whether a CAD PRS modifies the effect of statins on risk of acute coronary events within 10 years of statin initiation. IPTW is compared to linear regression with and without adjusting for baseline LDL cholesterol. The heading “modification of statin response by CAD PRS” reports an interaction between statin initiation and CAD PRS on risk of acute coronary events. Positive values of the interaction indicate that the CAD PRS is blunting the effect of statins, while negative values indicate it is conferring greater response.

**Extended Data Figure 4:**
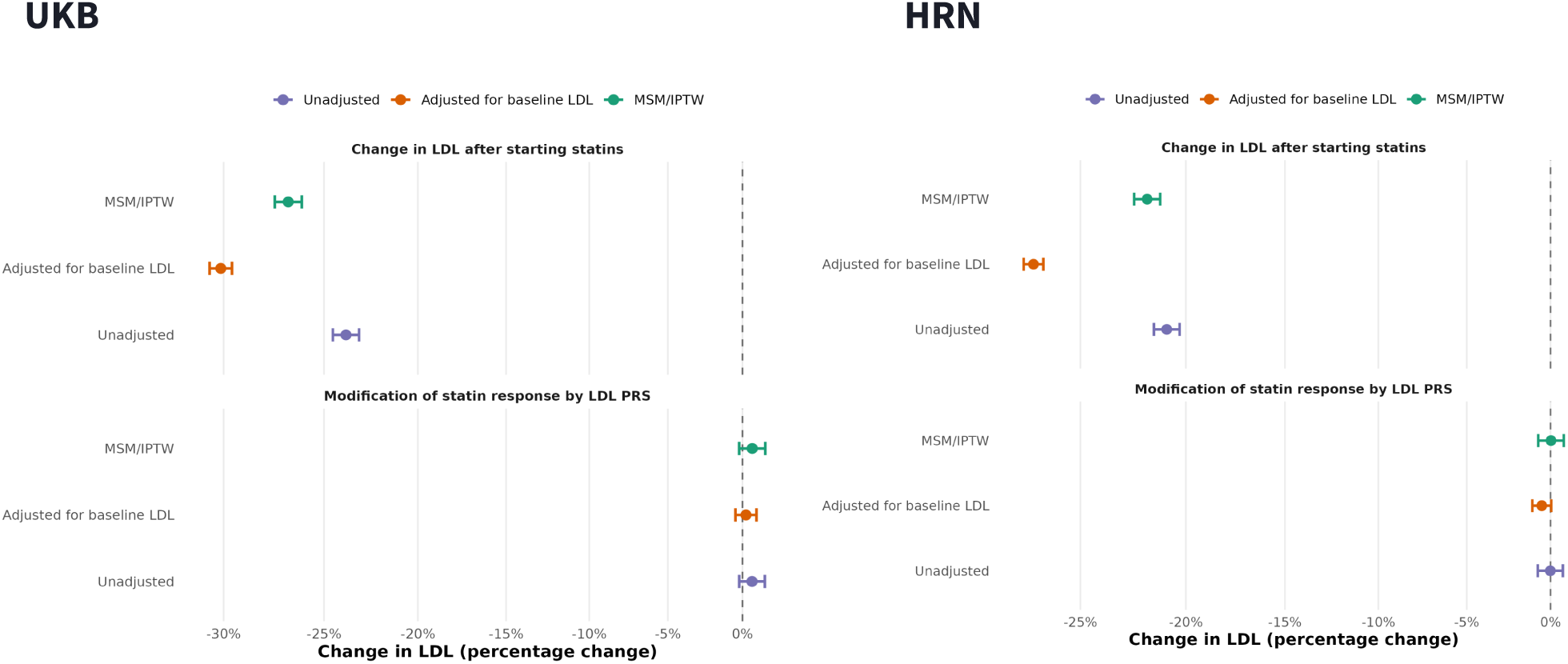
Primary analysis of UKB primary care and HRN electronic health records, testing whether a LDL PRS modifies the effect of statins on follow-up LDL cholesterol. IPTW is compared to linear regression with and without adjusting for baseline LDL cholesterol. The heading “modification of statin response by LDL PRS” reports an interaction between statin initiation and LDL PRS on follow-up LDL cholesterol. Positive values of the interaction indicate that the LDL PRS is blunting LDL response to statins, while negative values indicate it is conferring greater response.

**Extended Data Figure 5:**
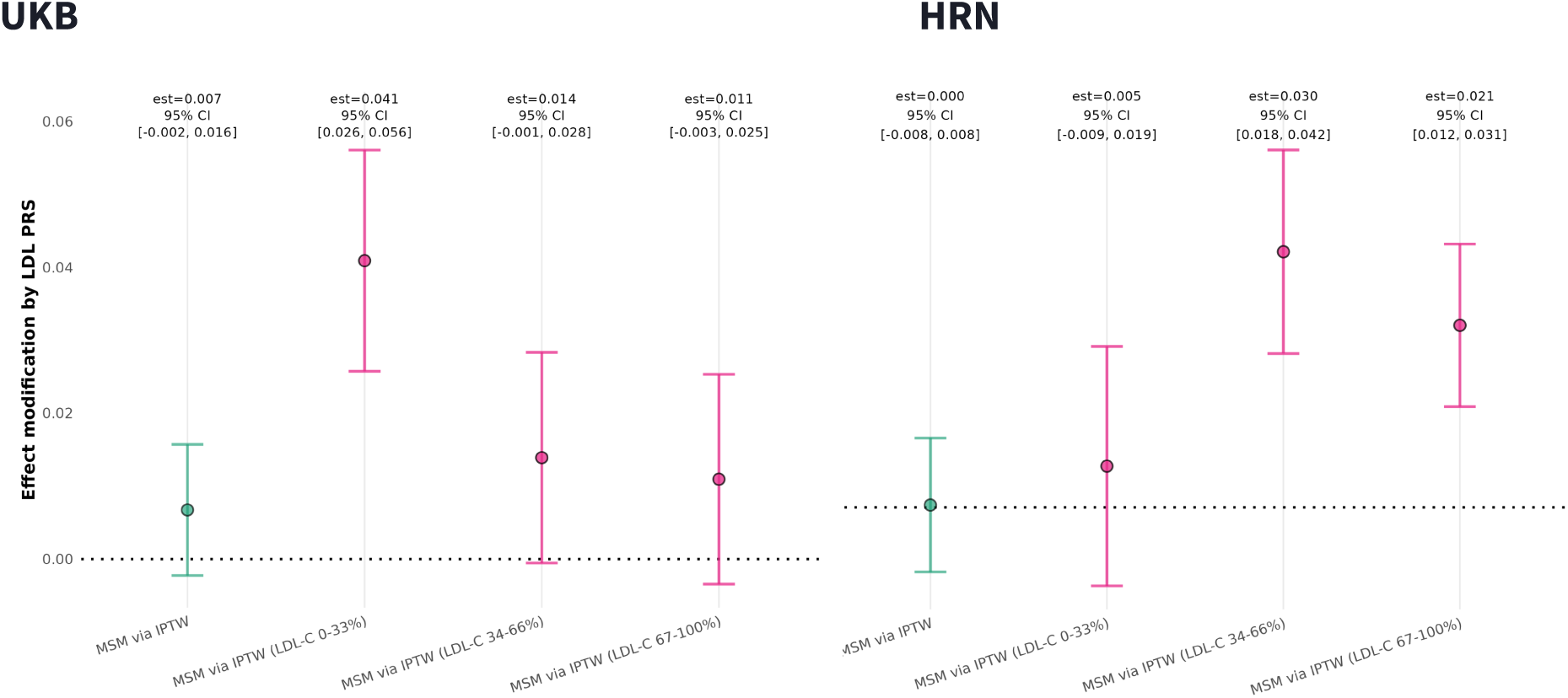
Cohort-specific estimates underlying the meta-analysis of LDL PRS-dependent statin response in Fig. 6. Interaction estimates between statin initiation and LDL PRS are shown separately for UK Biobank primary care (UKB; lek) and the Helix Research Network (HRN; right), in the full cohort and within each tertile of baseline LDL cholesterol. Within each analysis, inverse probability of treatment weighting was used to balance measured determinants of statin initiation between initiators and non-initiators. Positive interaction coefficients indicate that higher LDL PRS is associated with a blunted LDL cholesterol response to statin initiation; negative coefficients indicate greater response. Error bars show 95% confidence intervals.

## Supplementary Information

### Supplementary Note 1: Initiator-only change score designs may also biased for estimating PRS-dependent treatment response

Another common approach in pharmacoepidemiology is to restrict analyses to drug initiators and test whether polygenic risk predicts the change in an outcome after treatment. For example, studies of statin response may test whether an LDL cholesterol PRS is associated with the absolute or relative change in LDL cholesterol following treatment initiation^34–36^. These initiator-only designs are also susceptible to collider bias because treatment initiation depends on clinical characteristics, such as baseline LDL cholesterol, that are themselves influenced by polygenic risk. Restricting the analysis to statin initiators can therefore lead the PRS to associate with pre-versus-post change in LDL cholesterol, even if the PRS does not modify treatment response.

Collider bias is not the only challenge in interpreting these designs. Because initiator-only analyses do not compare treated with untreated individuals, they do not directly estimate whether the effect of treatment differs according to polygenic risk. Interpreting an association between PRS and pre-versus-post change in LDL cholesterol as treatment modification therefore requires strong assumptions: pre-treatment LDL cholesterol must fully capture the pathways through which polygenic risk influences both treatment initiation and post-treatment LDL cholesterol; the PRS must not influence post-treatment LDL cholesterol through other pathways; and there must be no other factors that affect both treatment initiation and post-treatment LDL cholesterol. Moreover, it is possible that changes observed after treatment initiation could have causes other than the treatment. Therefore, if LDL cholesterol would have changed over time even without treatment, change scores may also capture underlying temporal trends, regression to the mean, or measurement variation. Finally, their interpretation can depend on whether LDL cholesterol change is measured on an absolute or relative scale.

As illustrated in **Supplementary Fig. 1**, treatment initiation is influenced by both polygenic risk and other clinical risk factors. Restricting the analysis to people who initiate treatment can therefore create an association between the PRS and these other risk factors, even if they were otherwise independent. If those factors also influence post-treatment LDL cholesterol or modify statin response, the PRS can appear to predict LDL cholesterol change even when it does not truly modify the effect of treatment.

Since, as we outlined, there are many ways that change score analyses can go wrong, and our primary focus is on collider bias, we performed simulations under deliberately favourable conditions for the initiator-only change score design (**Methods; Table 3**). We assumed no time trends or regression to the mean, no direct effect of the PRS on post-treatment LDL cholesterol beyond its effect on pre-treatment LDL, and no direct effect of unmeasured factors on treatment initiation outside their influence through pre-treatment LDL.

We tested three analyses for the initiator-only change score analysis: relative change in LDL cholesterol both with and without adjusting for pre-treatment LDL, and the unadjusted absolute change in LDL cholesterol (**Supplementary Fig. 2**). Both the absolute change and adjusted relative change were biased in the simulation. The unadjusted relative change was unbiased, which we will discuss shortly in the real-data analysis.

We next ran the same three analyses in both HRN and UKB and meta-analyzed the estimates (**Supplementary Fig. 3**). LDL PRS was associated with apparent treatment response under all three specifications, but the direction and magnitude of association depended on how LDL cholesterol change was defined and whether pre-treatment LDL cholesterol was included as a covariate. Notably, a small negative association was observed for unadjusted relative change, while adjusted relative change produced a positive association. Thus, the choice of outcome scale and pre-treatment LDL cholesterol adjustment can produce opposite interpretations.

It is also notable that, although the simulation predicted that the LDL PRS should have no effect on unadjusted relative change in LDL cholesterol, the HRN and UKB meta-analysis found a small negative association (P = 0.007). There are several explanations consistent with this pattern. First, unmeasured factors may influence both statin initiation and post-treatment LDL cholesterol response; restricting the analysis to initiators can therefore induce associations between the LDL PRS and these factors that are not captured by the simulation. Second, repeated LDL cholesterol measurements are subject to regression to the mean. Because higher LDL PRS is associated with higher pre-treatment LDL cholesterol, mean regression of LDL cholesterol over time could produce a greater apparent relative reduction among individuals with higher PRS, even in the absence of PRS-dependent treatment response. These mechanisms are not mutually exclusive and both could contribute to the small negative association observed in HRN and UKB.

Taken together, these results suggest that initiator-only change score analyses should generally be avoided when studying polygenic modification of treatment response in observational cohorts. Study designs with an untreated comparison group are less susceptible to these forms of bias.

**Supplementary Figure 1:**
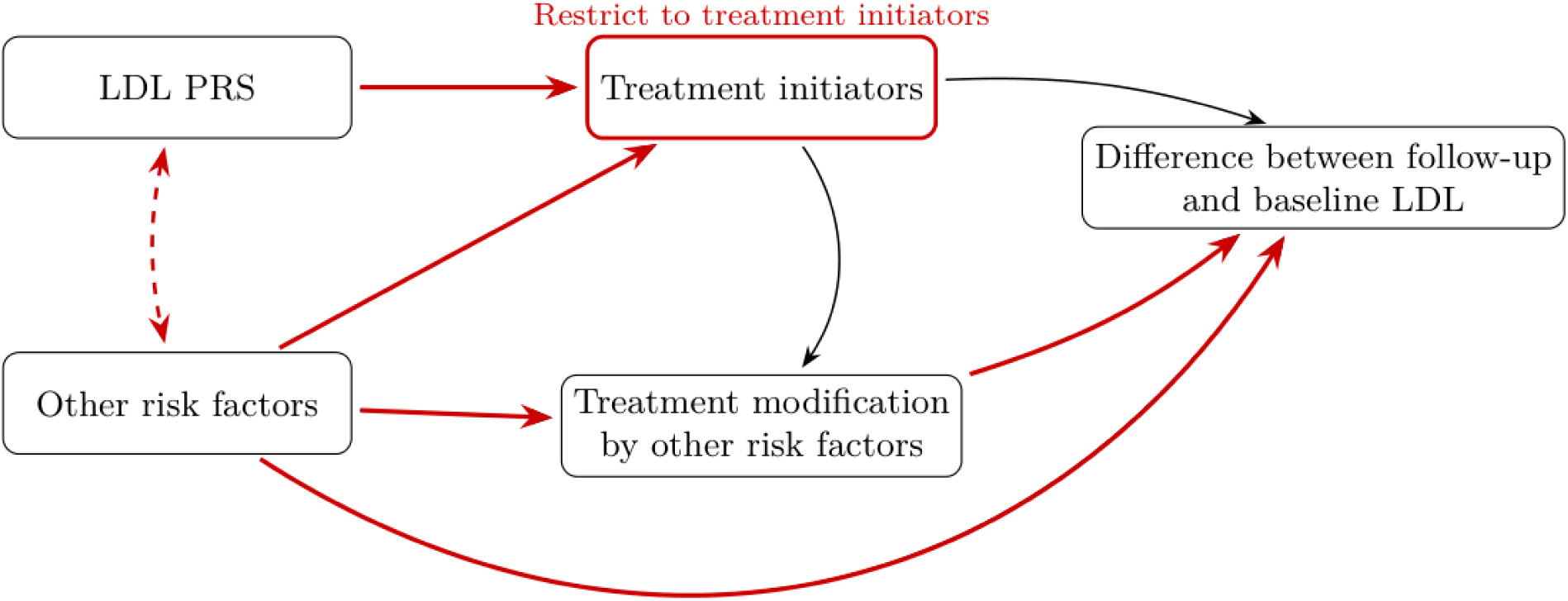
Collider bias in initiator-only analyses of PRS-dependent statin response. Restricting an analysis to treatment initiators conditions on a common consequence of LDL PRS and other clinical risk factors, inducing an association between them even if they were otherwise independent (dashed red arrow). If these other risk factors also influence LDL cholesterol change directly or modify statin response, LDL PRS can consequently appear to predict the difference between follow-up and baseline LDL cholesterol even when it does not modify the causal effect of treatment. Red arrows indicate pathways contributing to this bias.

**Supplementary Figure 2:**
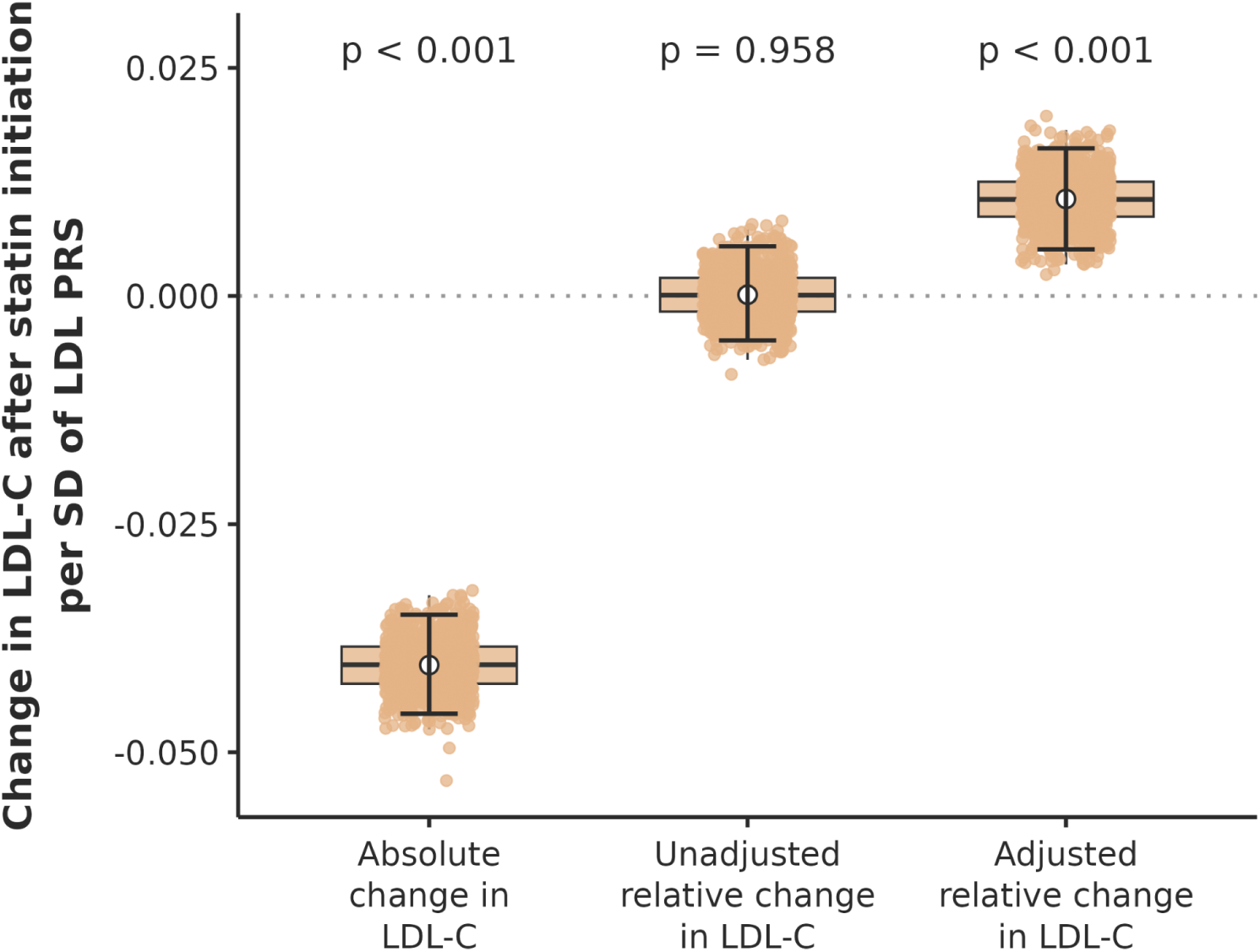
Initiator-only change-score analyses in simulation with no true PRS-dependent treatment response. We simulated 1,000 observational cohorts at the HRN sample size (n = 78,145) in which statin initiation depended on pre-treatment LDL cholesterol but the LDL PRS did not modify the causal effect of statins. Among statin initiators, we estimated the association of LDL PRS with absolute LDL cholesterol change, unadjusted relative change, and relative change adjusted for pre-treatment LDL cholesterol. Absolute change and baseline-adjusted relative change produced biased associations with LDL PRS, whereas unadjusted relative change was unbiased under the deliberately favourable assumptions of the simulation. Negative estimates indicate greater apparent LDL cholesterol reduction with increasing LDL PRS; positive estimates indicate a smaller apparent reduction.

**Supplementary Figure 3:**
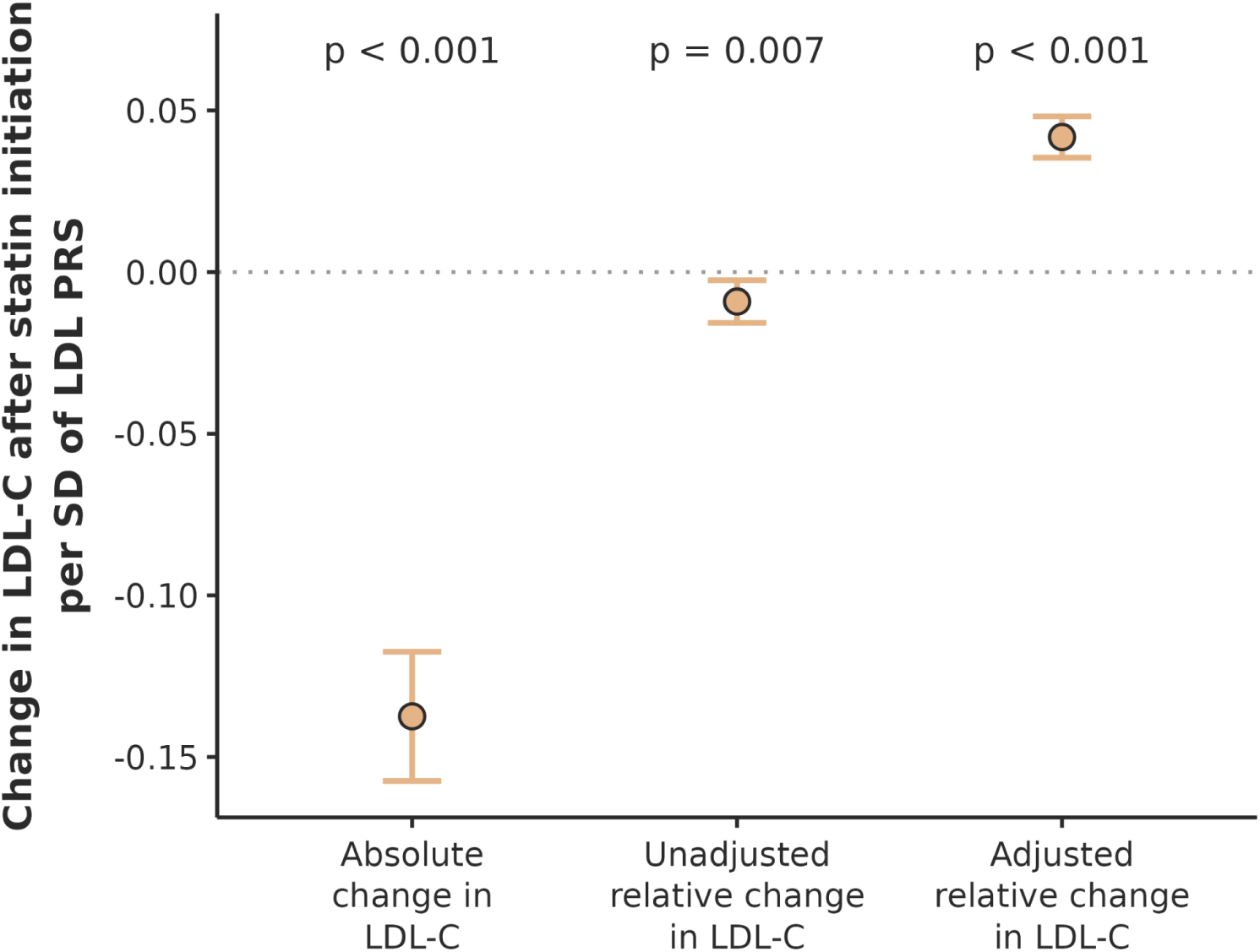
We applied the same three initiator-only analyses evaluated in simulation to statin initiators in UK Biobank (UKB) and the Helix Research Network (HRN), and combined cohort-specific estimates using inverse-variance weighting. LDL PRS was associated with apparent treatment response under all three specifications, but the direction and magnitude of association depended on how LDL cholesterol change was defined and whether pre-treatment LDL cholesterol was included as a covariate. Higher LDL PRS was associated with greater absolute LDL cholesterol reduction (pooled estimate −0.1374, 95% CI−0.1574 to −0.1174; P = 2.2 × 10⁻⁴¹), although this estimate showed substantial between-cohort heterogeneity (I² = 75%; Cochran’s Q = 3.95, P = 0.047). Unadjusted relative change showed a smaller negative association (−0.0091, 95% CI −0.0157 to −0.0025; P = 0.0068) with no evidence of heterogeneity (I² = 0%; Q = 0.44, P = 0.50). By contrast, relative change adjusted for pre-treatment LDL cholesterol produced a positive association (0.0418, 95% CI 0.0354 to 0.0482; P = 1.6 × 10⁻³⁷), again with no evidence of heterogeneity (I² = 0%; Q = 0.05, P = 0.82). Negative estimates indicate greater apparent LDL cholesterol reduction with increasing LDL PRS; positive estimates indicate a smaller apparent reduction.

